# Mathematical model describing CoViD-19 in São Paulo State, Brazil – Evaluating isolation as control mechanism and forecasting epidemiological scenarios of release

**DOI:** 10.1101/2020.04.29.20084830

**Authors:** Hyun Mo Yang, Luis Pedro Lombardi Junior, Fabio Fernandes Morato Castro, Ariana Campos Yang

## Abstract

We formulated a mathematical model considering young (below 60 years old) and elder (above 60 years) subpopulations to describe the introduction and dissemination of new coronavirus epidemics in the São Paulo State, Brazil. From the data collected in São Paulo State, we estimated the model parameters and calculated the basic reproduction number as *R*_0_ = 6.828. Considering isolation as a control mechanism, we varied the releasing proportions of young and elder persons to assess their epidemiological impacts. The best scenarios were release of young persons, but maintaining elder persons isolated. To avoid the collapse of the health care system, the isolation must be at least 80%.

## 1 Introduction

Mathematical models allow us to understand the progression of viral infections if the natural history of the disease is well documented. This understanding, in turn, permits forecasting epidemiological scenarios when control mechanisms are introduced aiming to reduce or eliminate the infection. In the case of coronavirus disease 2019 (CoViD-19), which is caused by severe acute respiratory syndrome coronavirus 2 (SARS-CoV-2), a strain of the RNA-based SARS-CoV-1, the possibility of obtaining scenarios is of fundamental importance. The reason is that in serious cases due to SARS-CoV-2 (new coronavirus) infection, immune cells overreact and attack the lung cell causing acute respiratory disease syndrome and possibly death. In general, the fatality rate in elder patients (60 years or more) is much higher than the average, and under 40 years seems to be around 0.2%. [10].

Due to the rapid spreading out of new coronavirus, its distribution is currently worldwide (pandemic). This virus can be transmitted by droplets that escape the lungs through coughing or sneezing and infect humans (direct transmission), or they are deposited in surfaces and infect humans when in contact with this contaminated surface (indirect transmission). The virus enters into susceptible persons through the nose, mouth, or eyes, and infects cells in the respiratory tract, being capable to release millions of new viruses. Like all RNA-based viruses, the new coronavirus tends to mutate faster than DNA-viruses, but lower than influenza viruses

Currently, there is not a vaccine, neither efficient treatment, although many drugs (chloroquine, for instance) are under clinical trial. Hence, isolation is the main, if not unique, way of controlling the dissemination of this virus in a population aiming the change in the natural history of disease propagation (this change is commonly known as the flattening curve of epidemics). Nevertheless, this isolation arises an important question: are there reliable strategies to release these isolated persons aiming to avoid the retaken of its original progression of infection?

Many mathematical and computational models are being used to describe current new coronavirus pandemics. In the mathematical models, there is a fundamental threshold (see [2]) called the basic reproduction number, which is defined as the secondary cases produced by one case introduced in a completely susceptible population, which is denoted by *R*_0_. When a control mechanism is introduced, this number is reduced and is called as the reduced reproduction number *R_r_*. Ferguson *et al*. [5] proposed a model to investigate the effects of isolation of susceptible persons. They analyzed two scenarios, called by them as mitigation and suppression, and predicted the numbers of severe cases and deaths due to CoViD-19 without control mechanism, and compared them with those numbers when control is introduced.

In this paper, we formulate a mathematical model based on ordinary differential equations, aiming the understanding of the dynamics (or trajectories of dissemination) of new coronavirus transmission, and applying this model to forecast changes in the dynamics under intervention. The model considers pulse isolation and a series of pulses of release (see [11] for a series of pulses vaccination). The model aims to describe the onset and subsequent spread of the new coronavirus in São Paulo State, Brazil. The understanding of the dynamics means that the model parameters are fitted against data collected in São Paulo State, Brazil. These estimated parameters are then used to study the potential scenarios when an intervention is introduced as a control mechanism. São Paulo State adopted the isolation of persons as the controlling mechanism, which can not be isolated indefinitely. Our main aim is to obtain epidemiological scenarios when releasing strategies will be implemented after isolation.

The paper is structured as follows. In Section 2, we introduce a model, which is numerically studied in Section 3. Discussions are presented in Section 4, and conclusions in Section 5.

## 2 Material and methods

In a community where SARS-CoV-2 (new coronavirus) is circulating, the risk of infection is greater in elder than young persons, as well as under increased probability of being symptomatic and higher CoViD-19 induced mortality. Hence, the community is divided into two groups, composed by young (under 60 years old, denoted by subscript *y*), and elder (above 60 years old, denoted by subscript *o*) persons. The vital dynamics of this community are described by per-capita rates of birth (*ϕ*) and mortality (*µ*).

For each sub-population *j* (*j = y, o*), all persons are divided into seven classes: susceptible *S_j_*, susceptible persons who are isolated *Q_j_*, exposed *E_j_*, asymptomatic *A_j_*, asymptomatic persons who are caught by test and then isolated *Q*_1_*_j_*, symptomatic persons at initial phase of CoViD-19 (or pre-diseased) *D*_1_*_j_*, pre-diseased persons caught by test and then isolated, plus mild CoViD-19 (or non-hospitalized) *Q*_2_*_j_*, and symptomatic persons with severe CoViD-19 (hospitalized) *D*_2_*_j_*. However, all young and elder persons in classes *A_j_*, *Q*_2_*_j_*, and *D*_2_*_j_* enter into the same immune class *I*.

The natural history of new coronavirus infection is the same for young (*j = y*) and elder (*j = o*) subpopulations. We assume that only persons in the asymptomatic (*A_j_*) and pre-diseased (*D*_1_*_j_*) classes are transmitting the virus, and other infected classes (*Q*_1_*_j_*, *Q*_2_*_j_* and *D*_2_*_j_*) are under voluntary or forced isolation. Susceptible persons are infected according to *λ_j_S_j_* (known as mass action law [2]) and enter into classes *E_j_*, where *λ_j_* is the per-capita incidence rate (or force of infection) defined by *λ_j =_ λ* (*δ_jy_* + *ψδ_jo_*), with *λ* being

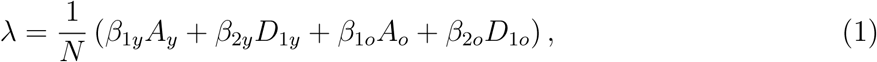

where *δ_ij_* is Kronecker delta, with *δ_ij =_* 1 if *i = j*, and 0, if *i* ≠ *j*; and *β*_1_*_j_* and *β*_2_*_j_* are the transmission rates, that is, the rates at which a virus encounters a susceptible people and infects. After an average period 1*/σ_j_* in class *E_j_*, where *σ_j_* is the incubation rate, exposed persons enter into the asymptomatic *A_j_* (with probability *p_j_*) or pre-diseased *D*_1_*_j_* (with probability 1 − *p_j_*) classes. After an average period 1*/γ_j_* in class *A_j_*, where *γ_j_* is the infection rate of asymptomatic persons, symptomatic persons acquire immunity (recovered) and enter into immune class *I*. Another route of exit from class *A_j_* is being caught by a test at a rate *η_j_* and enters into class *Q*_1_*_j_* and, then, after a period 1*/γ_j_*, enters into class *I*. Possibly asymptomatic persons are in voluntary isolation, which is described by the voluntary isolation rate *χ_j_*. With respect to symptomatic persons, after an average period 1*/γ*_1_*_j_* in class *D*_1_*_j_*, where *γ*_1_*_j_* is the infection rate of pre-diseased persons, pre-diseased persons enter into non-hospitalized *Q*_2_*_j_* (with probability *m_j_*) or hospitalized *D*_2_*_j_* (with probability 1 − *m_j_*) class. Hospitalized persons acquire immunity after a period 1*/γ*_2_*_j_*, where *γ*_2_*_j_* is the recovery rate of severe CoViD-19, and enter into immune class *I* or die under the disease induced (additional) mortality rate *α_j_*. Another route of exiting *D*_2_*_j_* is by treatment, described by the treatment rate *θ_j_*. After an average period 1*/γ_j_* in class *Q*_2_*_j_*, non-hospitalized persons acquire immunity and enter into immune class *I*, or enter into class *D*_2_*_j_* at a relapsing rate *ξ_j_*.

In the model, we consider pulse isolation and intermittent (series of pulses) release of persons. We assume that there is a unique pulse in isolation at time 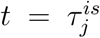, described by 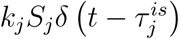, but there are *m* intermittent releases described by 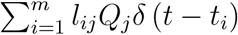, where 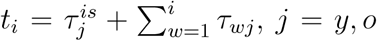, and *δ* (*x*) is Dirac delta function, that is, *δ* (*x*)= ∞, if *x =* 0, otherwise, *δ* (*x*) = 0, with 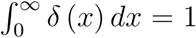. The parameters *k_j_* and *l_ij_*, *i =* 1, 2, ···,*m*, are the fractions of *i*-th release of isolated persons, and *τ_wj_* is the period between successive releases.

Figure 1 shows the flowchart of the new coronavirus transmission model.

**Figure 1:**
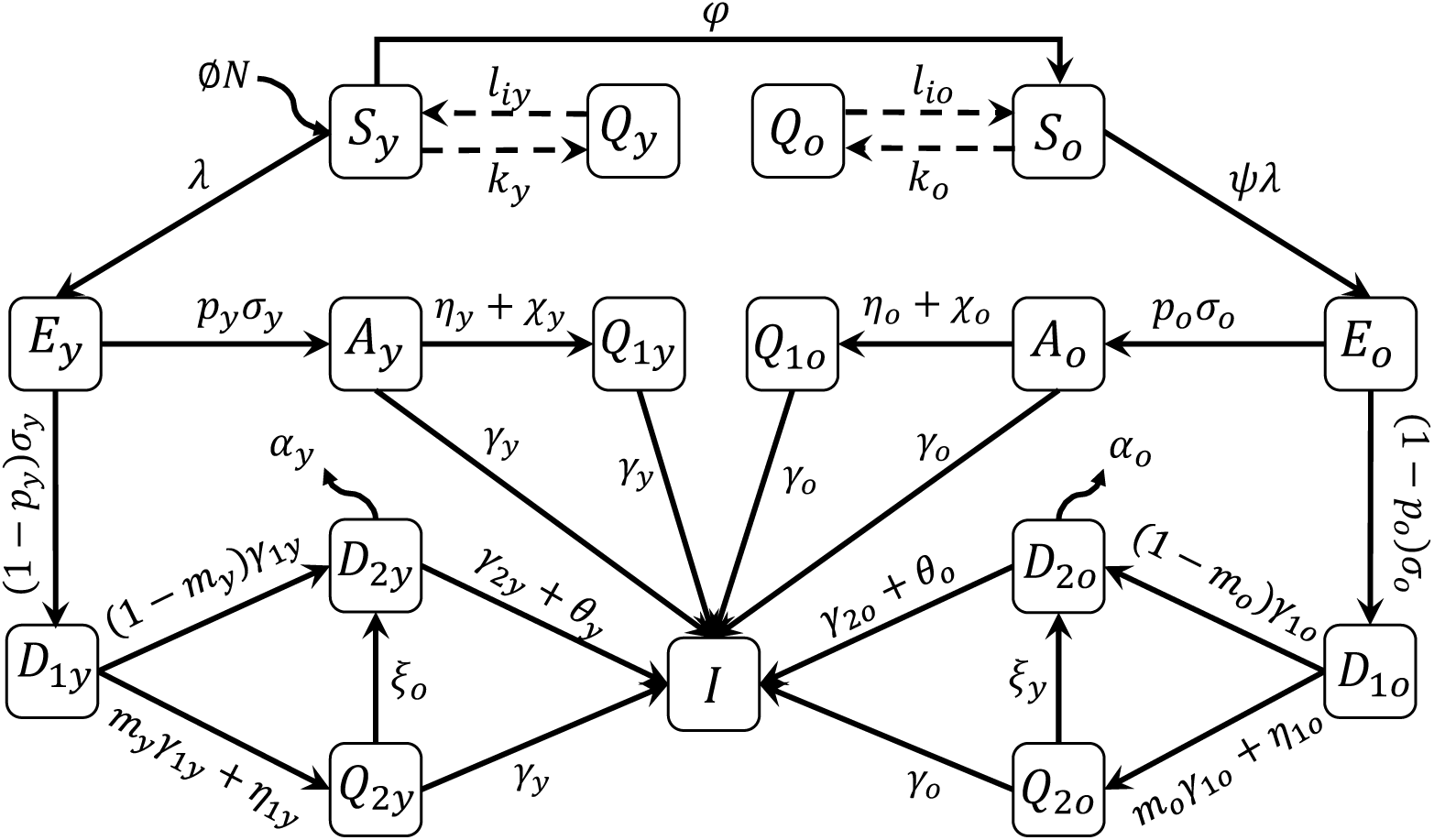
The flowchart of new coronavirus transmission model with variables and parameters.

The new coronavirus transmission model, based on above descriptions summarized in Figure 1, is described by system of ordinary differential equations, with *j = y, o*. Equations for susceptible persons are

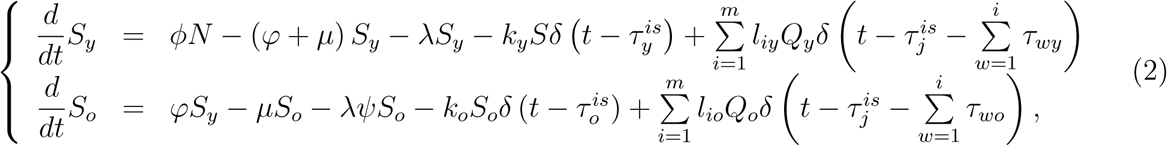

for infectious persons,

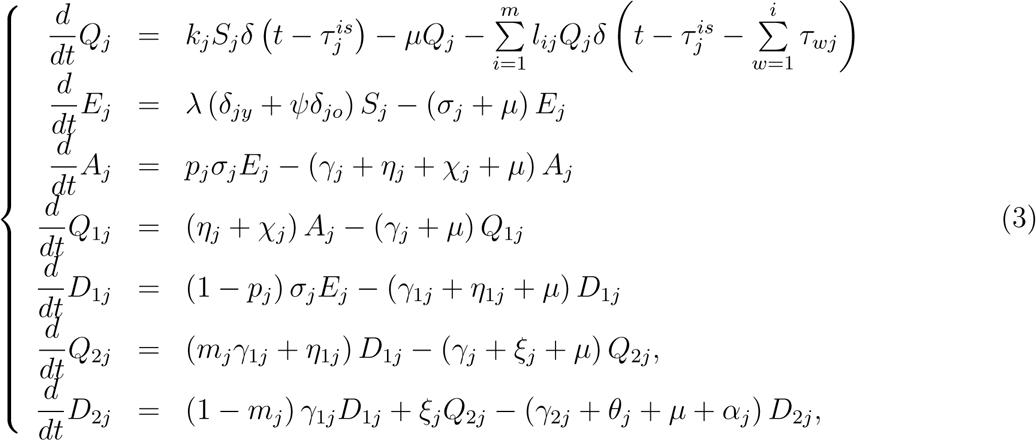

and for immune persons,

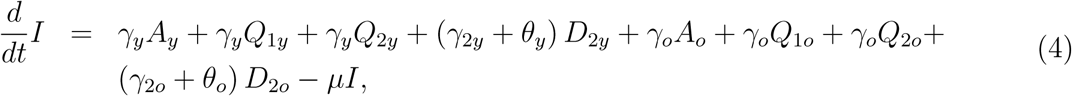

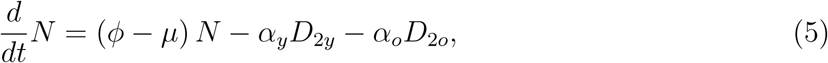

with the initial number of population at *t =* 0 being *N*(0) = *N*_0 =_ *N*_0_*_y_* + *N*_0_*_o_*, where *N*_0_*_y_* and *N*_0_*_o_* are the number of young and elder persons at *t =* 0. If *ϕ = µ* +(*α_y_D*_2_*_y_* + *α_o_D*_2_*_o_*) */N*, the total size of the population is constant.

Table 1 summarizes the model variables.

**Table 1:** Summary of the model variables (*j = y, o*).

| Symbol | Meaning |
| --- | --- |
| $S_j$ | Susceptible persons |
| $Q_j$ | Isolated among susceptible persons |
| $E_j$ | Exposed with new coronavirus persons |
| $A_j$ | Asymptomatic persons |
| $Q_{1j}$ | Isolated among asymptomatic persons by test |
| $D_{1j}$ | Initial symptomatic (pre-diseased) persons |
| $Q_{2j}$ | Isolated among pre-diseased persons by test |
| $D_{2j}$ | Severe CoViD-19 persons |
| $I_j$ | Immune (recovered) persons |

The non-autonomous system of equations (2), (3), and (4) is simulated letting intermittent interventions to the initial and boundary conditions. Hence, the equations for susceptible and isolated persons become

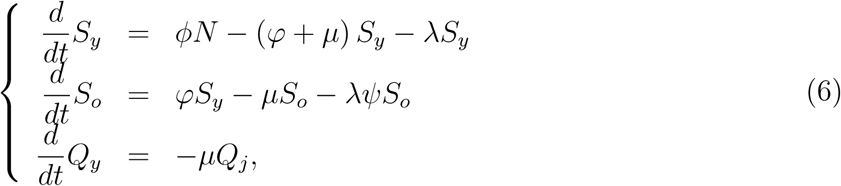

*j = y, o*, and other equations are the same.

For the system of equations (6), (3), and (4), the initial conditions (at *t =* 0) are, for *j = y, o*,

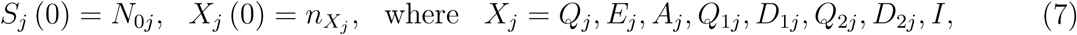

and 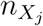 is a non-negative number. For instance, 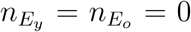 means that there is not any exposed individual (young and elder) at the beginning of epidemics. We split the boundary conditions into isolation and release, and assume that 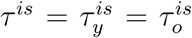 and *τ_i =_ τ_iy =_ τ_io_*, for *i =* 1, 2, ···, *m*, then 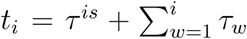 A unique isolation at *t = τ^is^* is described by the boundary conditions

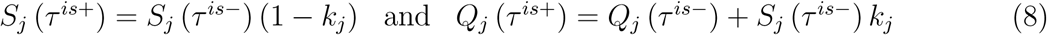

plus

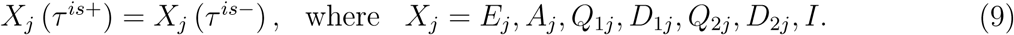

where we have 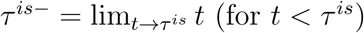, and 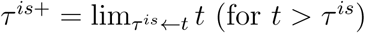. The boundary conditions for a series of pulses released at 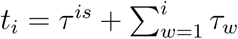, for *i =* 1, 2, ···, *m*, are

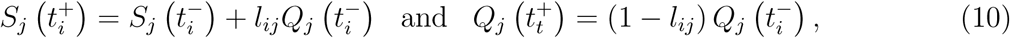

plus

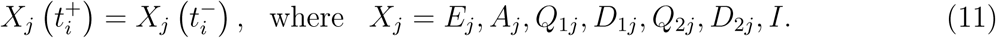

If *τ = τ_i_*, then *t_i =_ τ^is^*+ *iτ*. If isolation is applied to a completely susceptible population, at *t =* 0, there are not any infectious person, so *S*(0) = *N*_0_. If isolation is done at 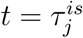 without screening of persons harboring the virus, then many of them could be isolated with susceptible persons.

Table 2 summarizes the model parameters and values (for elder classes, values are between parentheses).

**Table 2:** Summary of the model parameters (*j = y, o*) and values (rates in *days*^−1^, time in *days* and proportions are dimensionless). Some values are calculated (_&_), or varied (_#_), or assumed (^*^), or estimated (^**^) or not available (^***^).

| Symbol | Meaning | Value |
| --- | --- | --- |
| $\mu$ | Natural mortality rate | $1/(75 \times 360)[8]$ |
| $\phi$ | Birth rate | $1/(75 \times 360)^*$ |
| $\varphi$ | Aging rate | $6.7 \times 10^{-6}$ |
| $\sigma_y (\sigma_o)$ | Incubation rate | $1/5 (1/5)[10]$ |
| $\gamma_y (\gamma_o)$ | Infection rate of asymptomatic persons | $1/10 (1/11)[10]$ |
| $\gamma_{1y} (\gamma_{1o})$ | Infection rate of pre-diseased persons | $1/4 (1/4)[10]$ |
| $\gamma_{2y} (\gamma_{2o})$ | Recovery rate of severe CoViD-19 | $1/10 (1/14)[10]$ |
| $\xi_y (\xi_{yo})$ | Relapsing rate of pre-diseased persons | $0 (0)^*$ |
| $\alpha_y (\alpha_o)$ | Additional mortality rate | $0.0009 (0.009)^{**}$ |
| $\eta_y (\eta_o)$ | Testing rate among asymptomatic persons | $0 (0)^{***}$ |
| $\chi_y (\chi_o)$ | Voluntary isolation rate of asymptomatic persons | $0 (0)^{***}$ |
| $\eta_{1y} (\eta_{1o})$ | Testing rate among pre-diseased persons | $0 (0)^{***}$ |
| $k_y (k_o)$ | Proportion of Isolated susceptible persons | $0 (0)^*$ |
| $l_{iy} (l_{io})$ | Proportion released at time $t_i$ | $0 (0)^*$ |
| $\tau^{is}$ | Time of the introduction of isolation | 27 |
| $\tau_1 (\tau_i)$ | Time of first ($i$-th) releasing | $72(10)^*$ |
| $\theta_y (\theta_o)$ | Treatment rate | $0(0)^{***}$ |
| $\beta_{1y} (\beta_{1o})$ | Transmission rate due to asymptomatic persons | $0.76 (0.76)^{**}$ |
| $\beta_{2y} (\beta_{2o})$ | Transmission rate due to pre-diseased persons | $0.76 (0.76)^{**}$ |
| $\psi$ | Scaling factor of transmission among elder persons | $1.17^{**}$ |
| $p_y (p_o)$ | Proportion of asymptomatic persons | $0.8(0.75)^*$ |
| $m_y (m_o)$ | Proportion of mild (non-hospitalized) CoViD-19 | $0.8 (0.75)[3]$ |

From the system of equations (2), (3), and (4) we can derive some epidemiological parameters: new cases, new CoViD-19 cases, severe CoViD-19 cases, number of deaths due to CoViD-19, isolated persons, and number of occupied beds in hospitals.

The number of persons infected with new coronavirus is given by *E_y_* + *A_y_* + *Q*_1_*_y_* + *D*_1_*_y_* + *Q*_2_*_y_* + *D*_2_*_y_* for young persons, and *E_o_* + *A_o_* + *Q*_1_*_o_* + *D*_1_*_o_* + *Q*_2_*_o_* + *D*_2_*_o_* for elder persons. The incidence rates are

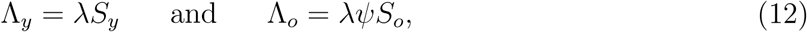

where the per-capita incidence rate *λ* is given by equation (1), and the numbers of new cases *C_y_* and *C_o_* are

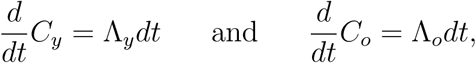

with *C_y_*(0) = 0 and *C_o_*(0) = 0

The number of CoViD-19 cases Δ*_y_* and Δ*_o_* are given by exist from *A_y_, D*_1_*_y_, A_o_*, and *D*1*o*, that is,

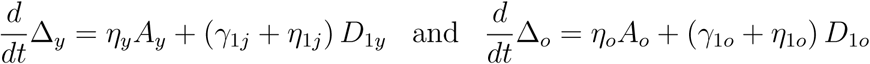

with Δ*_y_*(0) = 0 and Δ*_o_*(0) = 0, which are entering in classes *Q*_1_*_y_*, *D*_2_*_y_*, *Q*_2_*_y_*, *Q*_1_*_o_*, *D*_2_*_o_*, and *Q*_2_*_o_*.

The numbers of severe CoViD-19 (hospitalized) cases Ω*_y_* and Ω*_o_* are given by exits from *D*_1_*_y_*, *Q*_2_*_o_*, *D*_2_*_o_*, and *Q*_2_*_y_*, that is,

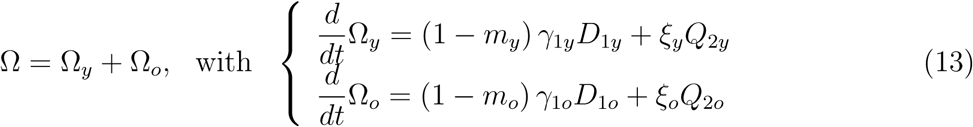

with Ω*_y_*(0) = Ω*_o_*(0) = 0, which are entering in classes *D*_2_*_y_* and *D*_2_*_o_* at each day.

The number of deaths caused by severe CoViD-19 cases Π can be calculated from hospitalized cases. This number of deaths is

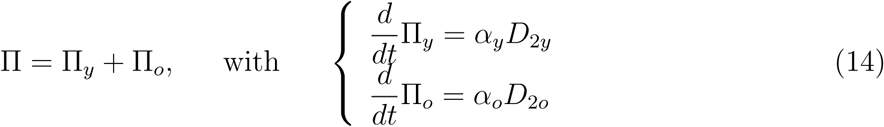

with Π*_y_*(0) = Π*_o_*(0) = 0.

The number of susceptible persons in isolation in the absence of release is obtained from

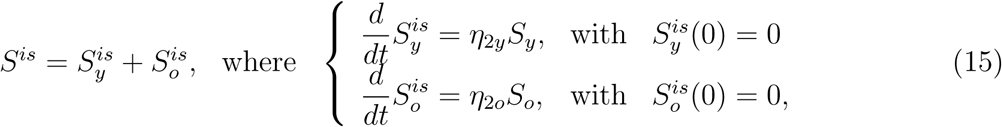

where the corresponding fractions of isolated susceptible persons are 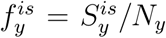 and 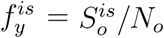

Finally, the number of beds *B* occupied during the evolving of epidemics is, for *j = y, o*,

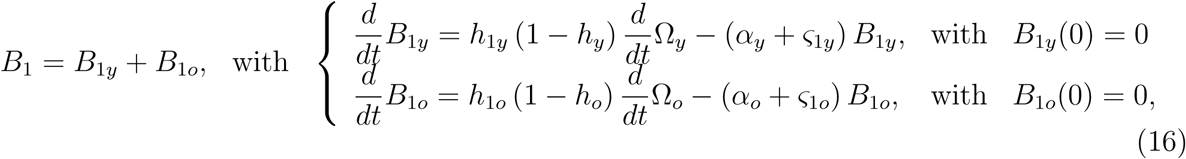

for beds in hospitals, and

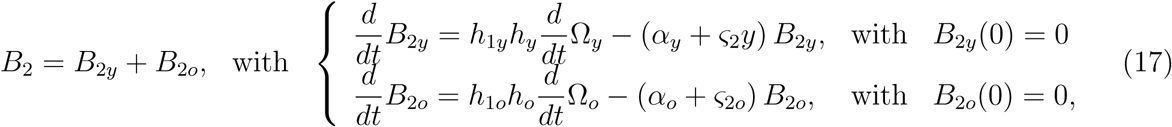

for beds in ICU. The fraction of severe CoViD-19 needing ICU is *h_j_*, and 1*/ς*_1_*_j_* and 1*/ς*_2_*_j_* are the average occupying time of beds in hospital and ICU, equal for young and elder persons, where *ς*_1_*_j_* and *ς*_2_*_j_* are the discharging rates from hospital and ICU, for *j = y, o*. The fraction *h*_1_*_j_* is severe CoViD-19 needing prolonged hospital care. The total number of occupied beds is *B = B*_1_ + *B*_2_.

The system of equations (2), (3), and (4) is non-autonomous. Nevertheless, the fractions of persons in each compartment approach the steady state (see Appendix A). Hence, at *t =* 0, the basic reproduction number *R*_0_ is obtained substituting 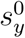 and 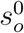 by 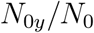 and 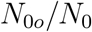 in equation (A.4), resulting in

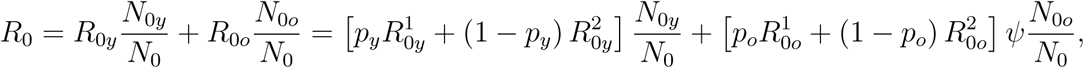

using equations (A.8) and (A.9).

Let us use the approximated effective reproduction number *R_ef_* given by equation (A.11), that is, *R_ef =_ R*_0_ (*S_y_/N* + *S_o_/N*). For *t>* 0, we have the effective reproduction number *R_ef_*, with *R_ef_* (0) = *R*_0_ at *t =* 0, which decreases as susceptible persons decrease. However, at *t = τ^is^* a pulse in isolation is introduced, hence we have *R_ef_* (*τ^is^*^+^) = *R_r_*, where the reduced reproduction number R*_r_* is given by

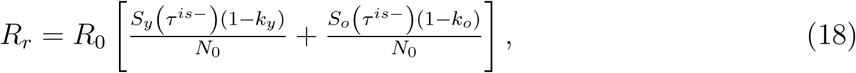

and *S_y_* (*τ^is^*^−^) and *S_o_* (*τ^is^*^−^) are the number of susceptible young and elder persons at the time just before the introduction of isolation. Notice that at *t = τ^is^*, R*_ef_* (*τ^is^*^−^) jumped down to *R_ef_* (*τ^is^*^+^). At *i*-th release time *t_i_*, we have 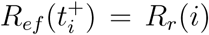, with the reduced reproduction number *R_r_*(*i*) being given by

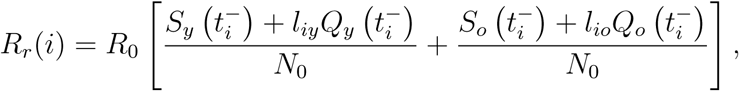

where 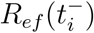 jumped up to 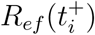 at *t = t_i_*. After *t>t_m_*, there is not release anymore, and *R_ef =_* 1 when *t* →∞, and the new coronavirus returns to the original dynamics driven by *R*_0_.

Given *N* and *R*_0_, let us evaluate the number of susceptible persons to trigger and maintain epidemics. Let us assume that all model parameters for young and elder classes and all transmission rates are equal, then *R*_0 =_ *σβ/* [(*σ* + *ϕ*)(*γ* + *ϕ*)] and *R_ef =_ R*_0_*S/N*, using the approximated *R_ef_* given by equation (A.11). Letting *R_ef =_* 1, the critical number of susceptible persons *S^th^* at equilibrium is

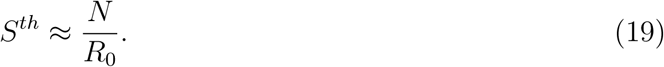

If *S >S^th^*, epidemics occurs and persists (*R_ef_ >* 1, non-trivial equilibrium point *P^*^*), and the fraction of susceptible individuals is *s^*^ =* 1*/R*_0_, where 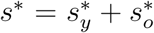; but if *S < S^th^*, epidemics occurs but fades out (*R_ef_ <* 1, trivial equilibrium point *P*^0^), and the fractions of susceptible individuals *s_y_* and *s_o_* at equilibrium are given by equation (A.4). Remember that the threshold *S^th^* is valid only if the pulse isolation introduced at *t =* 0 is maintained forever.

We apply the above results to study the introduction and establishment of the new coronavirus in São Paulo State, Brazil.

## 3 Results

The results obtained in the foregoing section are applied to describe the new coronavirus infections in São Paulo State, Brazil. The first confirmed case of CoViD-19, occurred on February 26, 2020, was a traveler returning from Italy on February 21, and being hospitalized on February 24. The first death due to CoViD-19 was a 62 years old male with comorbidity who never travelled to abroad, hence considered as autochthonous transmission. He manifested the first symptoms on March 10, was hospitalized on March 14, and died on March 16. On March 24, the São Paulo State authorities ordered isolation of persons acting in non-essential activities, as well as students of all levels until April 6, further isolation was extended to April 22, and postponed to May 10.

The dynamics of the new coronavirus propagation is obtained by evaluating the system of equations (2), (3), and (4) numerically using the 4*^th^* order Runge-Kutta method. Let us determine the initial conditions supplied to this system. In the São Paulo State, the number of inhabitants is *N* (0) = *N*_0 =_ 44.6×10^6^ according to SEADE [8]. The value of parameter *φ* given in Table 1 was calculated by the equation (A.4), *φ = bϕ/* (1 − *b*), where *b* is the proportion of elder persons. Using *b =* 0.153 in the São Paulo State [8], we obtained *φ =* 6.7 × 10^−6^ *days*^−1^, hence, 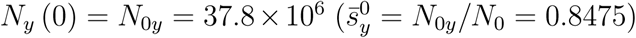 and 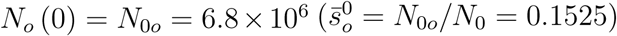. The initial conditions for susceptible persons are set to be *S_y_* (0) = *N*_0_*_y_* and *S_o_* (0) = *N*_0_*_o_*. For other variables, from Table 2, using *p_y =_* 0.8 and *m_y =_* 0.8, the ratio asymptomatic:symptomatic is 4 : 1, and the ratio mild:severe (non-hospitalized:hospitalized) CoViD-19 is 4 : 1. We also use these ratios for elder persons, even *p_o_* and *m_o_* are slightly lower. Hence, if we assume that there is one person in *D*_2_*_j_* (the first confirmed case), then there are 4 persons in *Q*_2_*_j_*. The sum (5) is the number of persons in class *D*_1_*_j_*, implying that there are 20 in class *A_j_*, hence, the sum (25) is the number of persons in class *E_j_*. Finally, we suppose that no one is isolated or tested, and immunized. (Probably the first confirmed COViD-19 person transmitted virus (since February 21 when returned infected from Italy), as well as other asymptomatic travelers returning from abroad.)

Therefore, the initial conditions supplied to the dynamic system (2), (3), and (4) are

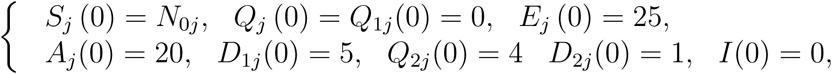

where the initial simulation time *t =* 0 corresponds to the calendar time February 26, 2020, when the first case was confirmed.

This section presents parameters estimation and epidemiological scenarios considering isolation as the control mechanism. In estimation and epidemiological scenarios, we assume that all transmission rates in young persons are equal, as well as in elder persons, that is, we assume that

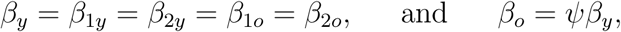

hence, the forces of infection are *λ_y =_* (*A_y_* + *D*_1_*_y_* + *A_o_* + *D*_1_*_o_*) *β_y_/N* and *λ_o =_ ψλ_y_*. The reason to include factor *ψ* is the reduced capacity of defense mechanism by elder persons (physical barrier, innate and adaptive immune responses, etc.). The force of infection takes into account all virus released by infectious individuals (*A_y_*, *D*_1_*_y_*, *A_o_*, and *D*_1_*_o_*), the rate of encounter with susceptible persons, and the capacity to infect them (see [12] [13]). Additionally, the amount aspired by susceptible persons can be determinant in the chance of infection and in the prognostic of CoViD-19 [19].

From data collected in the São Paulo State from February 26 until April 5, 2020, we fit transmission (*β_y_* and *β_o_*) and additional mortality rates (*α_y_* and *α_o_*), and the proportions of isolated persons (*k_y_* and *k_o_*) are estimated using data from March 24 until April 21. Once determined these parameters, we study potential scenarios introducing isolation as control mechanisms.

### 3.1 Parameters estimation

Reliable estimations of both transmission and additional mortality rates is crucial, aiming the prediction of new cases (to an adequate number of beds in a hospital, for instance) and deaths. When the estimation is based on a few number of data, that is, at the beginning of epidemics, some cautions must be taken, because the rates maybe over or under estimated. The reason is that in the very beginning phase of epidemic, the spreading out of infections and death increase exponentially without bound.

Currently, there is not a sufficient number of kits to detect infection by the new coronavirus. For this reason, test to confirm infection by this virus are done only in hospitalized persons, and also in persons who died, manifesting symptoms of CoViD-19. Hence, we have only data of hospitalized persons (*D*_2_*_y_* and *D*_2_*_o_*) and those who died (Π*_y_* and Π*_o_*). Taking into account hospitalized persons with CoViD-19, we fit the transmission rates, and for persons died due to CoViD-19, we fit the additional mortality rates. These rates are fitted applying the least square method (see [7]), that is

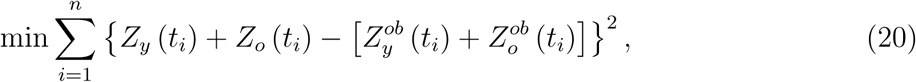

where min stands for the minimum value, *n* is the number of observations, *t_i_* is *i*-th observation time, *Z_j_* stands for *Ω_j_* given by equation (13), and Π*_j_* is given by equation (14); and 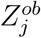 stands for the observed number of hospitalized persons 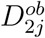 and number of died persons 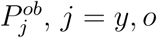 The fitted parameters are those minimizing the sum of squared differences.

Instead of using equation (20), the least square estimation method, we vary the parameters and choose better fitting by evaluating the sum of squared distances between curve and data.

#### 3.1.1 Fitting the transmission rates

The introduction of quarantine was at *t =* 27, corresponding to the calendar time March 24, but the effects are expected to appear later. Hence, we will estimate taking into account the confirmed cases from February 26 (*t =* 0) to April 5 (*t =* 39), hence *n =* 40 observations. It is expected that at around simulation time *t =* 36 (April 2), the effects of isolation will appear (the sum of incubation and pre-diseased infection periods (see Table 2) is around 9 days).

To estimate the transmission rates *β_y_* and *β_o_*, we let *α_y =_ α_o =_* 0 and the system of equations (2), (3), and (4), with initial conditions given by equation (7), is evaluated, and we calculate

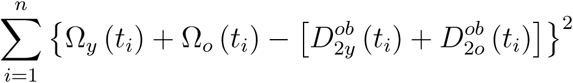

by varying *β_y_* and *β_o_*. We chose the transmission rates minimizing the sum of differences. Letting additional mortality rates equal to zero (*α_y =_ α_o =_* 0), we estimate *β_y_* and *β_o =_ ψβ_y_*, against hospitalized CoViD-19 cases (*D*_2 =_ *D*_2_*_y_* + *D*_2_*_o_*) data from the São Paulo State. The estimated values are *β_y =_* 0.75 and *β_o =_* 0.8775 (*days*^−1^), where Ψ = 1.17, resulting in the basic reproduction number *R*_0 =_ 6.828 (partials *R*_0_*_y =_* 5.587 and *R*_0_*_o =_* 1.241), according to equation (A.8). Figure 2(a) shows the estimated curve of *D*_2_ and observed data, plus two curves with lower transmission rates: *β_y =_* 0.55 and *β_o =_* 0.6435 (*days*^−1^), with *R*_0 =_ 5.008; and *β_y =_* 0.45 and *β_o =_* 0.5265 (*days*^−1^), with *R*_0 =_ 4.097 (partials *R*_0_*_y =_* 3.352 and *R*_0_*_o =_* 0.745). Figure 2(b) shows curves, from *t =* 30 until 180 of *D*_2_ for young, elder, and total persons for *R*_0 =_ 6.828 and 4.097.

**Figure 2:**
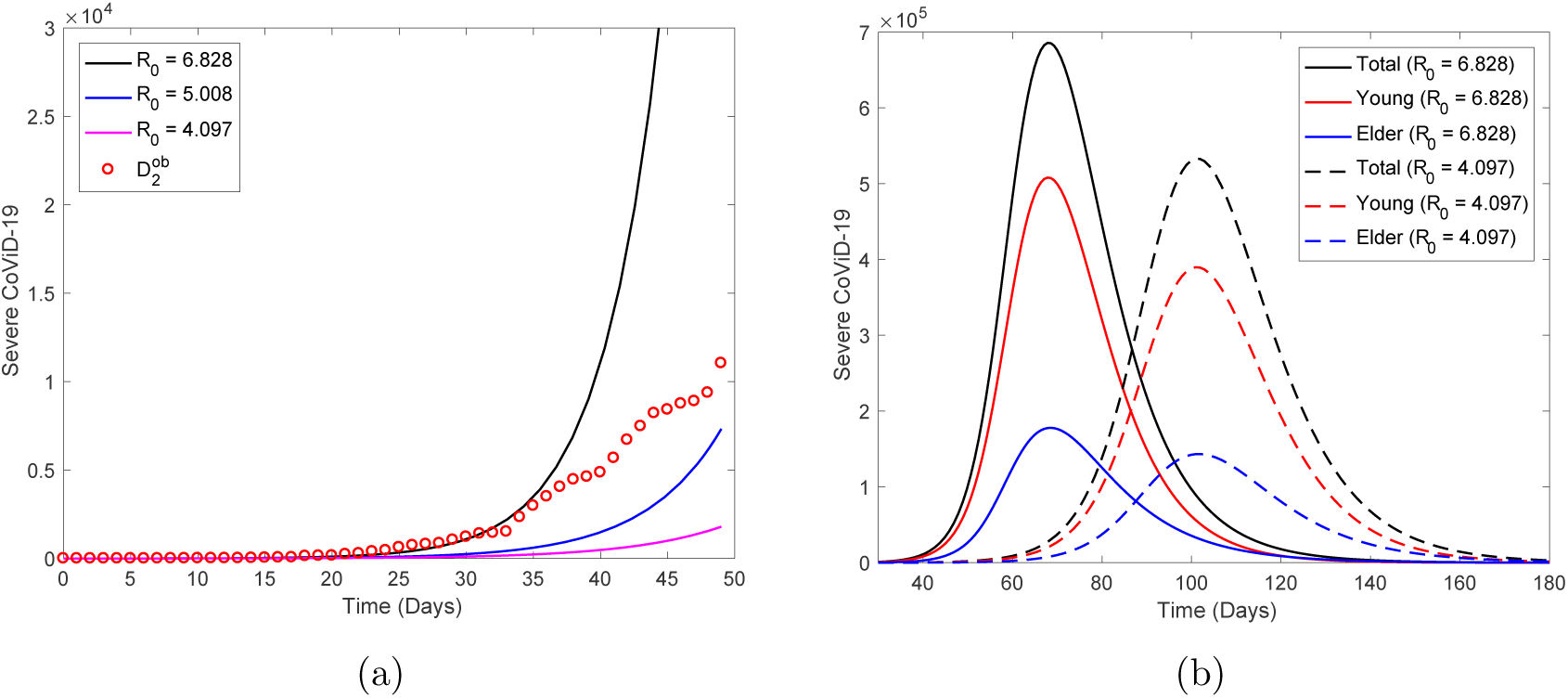
The estimated curve of severe CoViD-19 cases *D*_2_ (*β_y =_* 0.75 and *β_o =_* 0.8775 (*days*^−1^), with *R*_0 =_ 6.828) and observed data, plus two curves with lower transmission rates: *β_y =_* 0.55 and *β_o =_* 0.6435 (*days*^−1^), with *R*_0 =_ 5.008; and *β_y =_* 0.45 and *β_o =_* 0.5265 (*days*^−1^), with *R*_0 =_ 4.097. Estimation was done using data from *t =* 0 to *t =* 39 (a), from top to bottom as *R*_0_ increases, and the extended curves until *t =* 250 (b), from top to bottom for total, young and elder persons, with continuous (*R*_0 =_ 6.828) and dashed (*R*_0 =_ 4.097) curves.

From Figure 2(a), as *R*_0_ decreases, the estimations become worst. From Figure 2(b), the peak of higher *R*_0_ occurs at *t =* 68, while for lower *R*_0_, delayed in 33 days, at *t =* 101. The peaks for *R*_0 =_ 6.828 and 4.097 (between parentheses) are 6.86 × 10^5^ (5.33 × 10^5^), 5.08 × 10^5^ (3.898 × 10^5^) and 1.78 × 10^5^ (1.43 × 10^5^), respectively, for young, elder and total persons.

Let us estimate the critical number of susceptible persons *S^th^* from equation (19). For *R*_0 =_ 6.828, we have *S^th =^* 6.532 × 10^6^. Hence, for the São Paulo State, isolating 38.07 million (85.35%) or above persons is necessary to avoid the persistence of epidemics. The number of young persons is 0.27 million less than the threshold number of isolated persons to guarantee eradication of CoViD-19. For *R*_0 =_ 4.097, we have *S^th^ _=_* 10.87 × 10_6_, implying to isolate 33.73 million (75.63%) or above persons, which is 4.07 million less than young persons.

#### 3.1.2 Fitting the additional mortality rates

We estimate taking into account confirmed deaths from February 26 (*t =* 0) to April 5 (*t =* 39), hence *n =* 40 observations, as we did in the previous estimation.

To estimate the mortality rates α*_y_* and *α_o_*, we fix the previously estimated transmission rates *β_y_* and *β_o_* for *R*_0 =_ 6.828 and 4.097, and evaluate the system of equations (2), (3) and (4), with initial conditions given by equation (7), to calculate

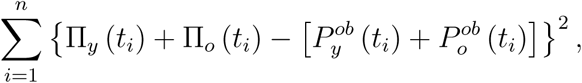

by varying *α_y_* and *α_o_*. We chose the mortality rates minimizing the sum of differences. Fixing the previously estimated transmission rates *β_y =_* 0.75 and *β_o =_* 0.8775 (both *days*^−1^), we estimate additional mortality rates *α_y_* and *α_o_* by varying *α_y_* and *α_o_*. From the fact that lethality among young persons is lower than elder persons, we let *α_y =_* 0.2*α_o_* [3], and estimate only one variable *α_o_*. The estimated rates are *α_y =_* 0.0052 and *α_o =_* 0.026 (*days*^−1^). For lower *R*_0_, fixing *β_y =_* 0.45 and *β_o =_* 0.5265 (*days*^−1^), we estimated *α_y =_* 0.08 and *α_o =_* 0.4(*days*^−1^). This is called the first estimation method.

The first estimation method used only one information: the risk of death is higher among elder than young persons (we used *α_y =_* 0.2*α_o_*). However, the lethality among hospitalized elder persons is around 10% [3]. Combining both findings, we assume that the deaths for young and elder persons are, respectively, 2% and 10% of accumulated cases when Ω*_y_* and Ω*_o_* approach plateaus (see Figures 4(a) and 5(a) below). This is called the second estimation method, which takes into account a second information besides the one used in the first estimation method. In this procedure, the estimated rates are *α_y =_* 0.0018 and *α_o =_* 0.009 (*days*^−1^) for both *R*_0_.

Figure 3 shows the estimated curves of Π = Π*_y_* +Π*_o_* provided by both methods of estimation and observed data for *R*_0 =_ 6.828 (a) and 4.097 (b). The second method of estimation fits very badly in the interval of estimation from *t =* 0 to 39.

**Figure 3:**
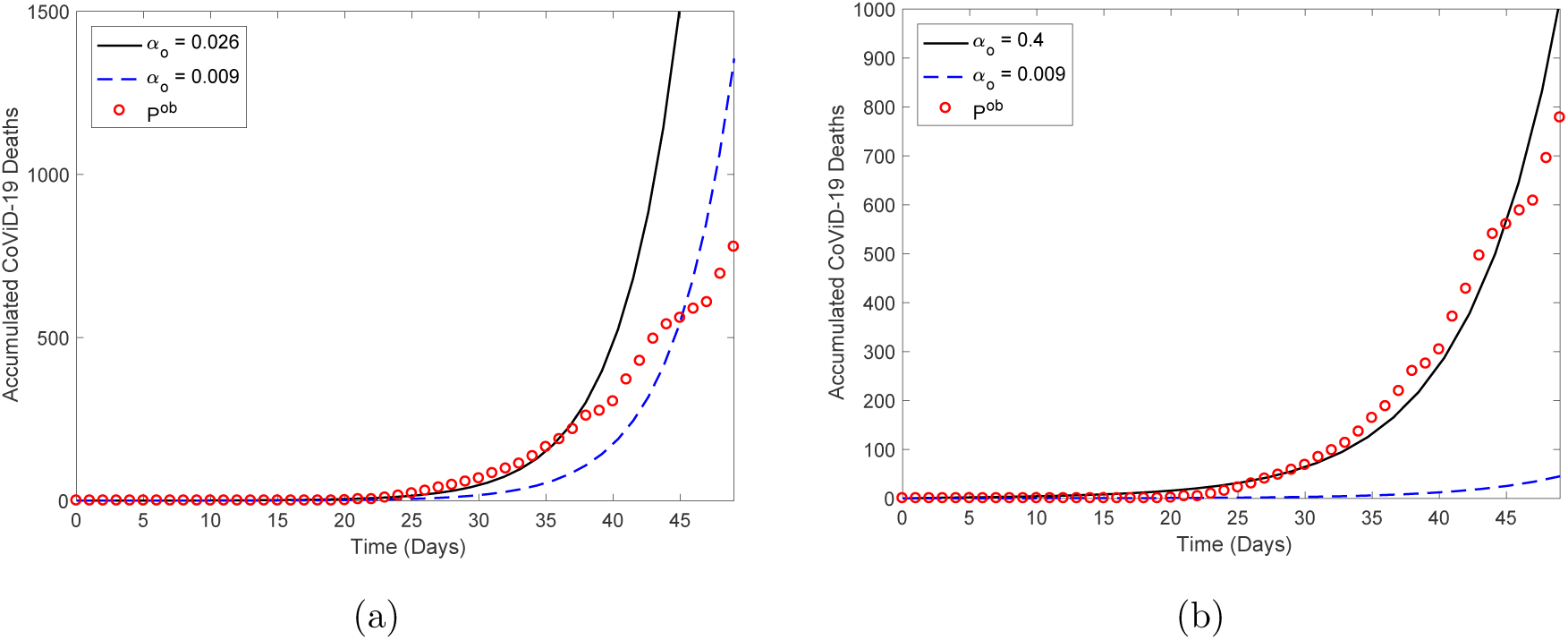
The estimated curves of Π provided by both methods of estimation and observed data from *t =* 0 until 39. The first estimation method provided for *R*_0 =_ 6.828, *α_y =_* 0.0052 and *α_o =_* 0.026 (a), and for *R*_0 =_ 4.097, *α_y =_* 0.08 and *α_o =_* 0.4 (b). Both figures show also the second method of estimation, which fits very badly in the interval of estimation, providing same *α_y =_* 0.0009 and *α_o =_* 0.009 (all in *days*^−1^).

For higher *R*_0 =_ 6.828, Figure 4(a) shows the estimated curves of accumulated number of severe CoViD-19 (Ω*_y_*, Ω*_o_*, and Ω = Ω*_y_* +Ω*_o_*), from equation (13). At *t =* 120 days, Ω approached an asymptote (or plateau), which can be understood as the time when the first wave of epidemics ends. The curves Ω*_y_*,Ω*_o_*, and Ω reach values at *t =* 200, respectively, 1.511 × 10^6,^0.426 × 10^6^, and 1.937 × 10^6^. Figure 4(b) shows the estimated curves of the accumulated number of CoViD-19 deaths (Π*_y_*,Π*_o_*, and Π = Π*_y_* +Π*_o_*), from equation (14). The values of Π*_y_*, Π*_o_*, and Π are, at *t =* 200, for the first method of estimation, respectively, 0.747 × 10^5^ (4.9%), 1.137 × 10^5^ (26.7%) and 1.883 × 10^5^ (9.72%), and for the second method of estimation, respectively, 2.67 × 10^4^ (1.77%), 4.767 × 10^4^ (11.19%) and 7.437 × 10^4^ (3.84%). The percentage between parentheses is the ratio Π*/*Ω.

**Figure 4:**
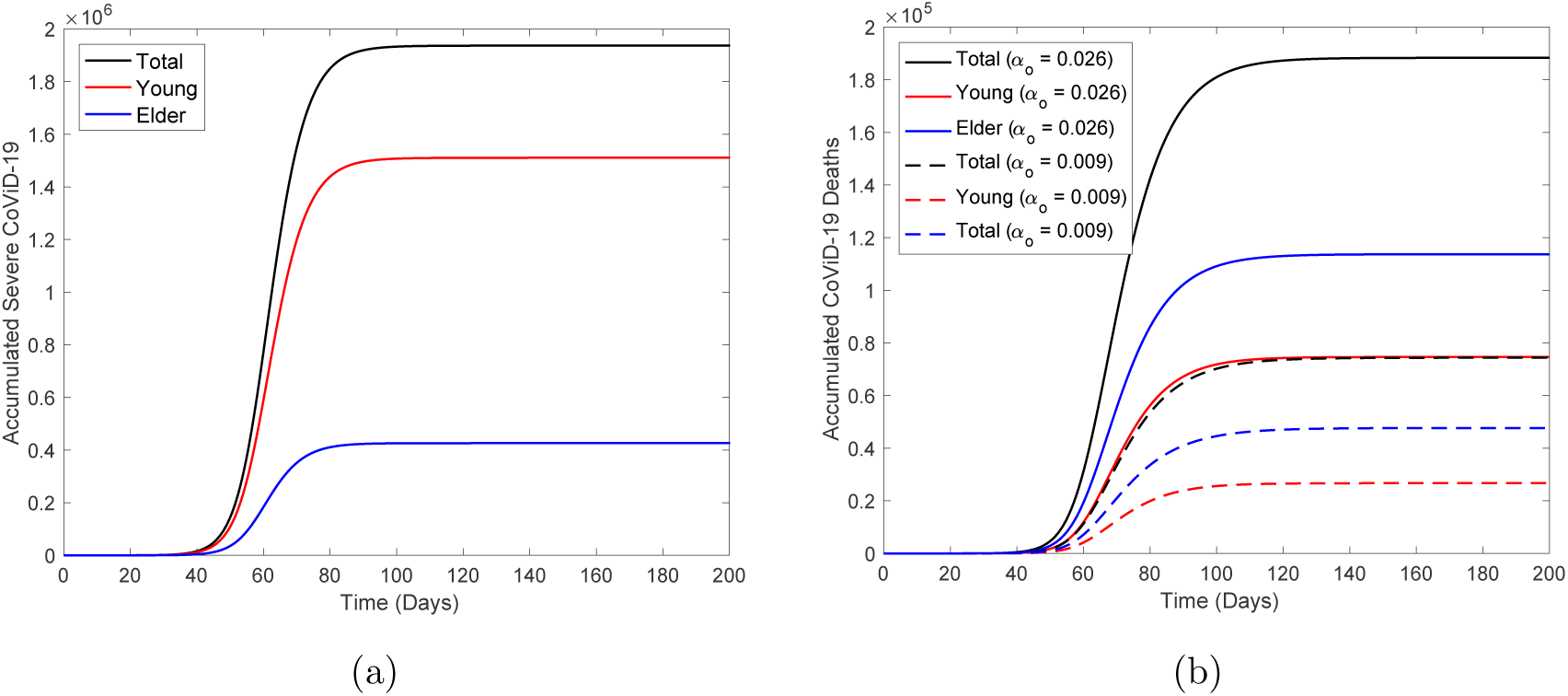
For *R*_0 =_ 6.828, the estimated curves of accumulated number of severe CoViD-19 (Ω*_y_*, Ω*_o_* and Ω = Ω*_y_* +Ω*_o_*) (a), from top to bottom for total, young and elder persons, and accumulated number of CoViD-19 deaths (Π*_y_*, Π*_o_* and Π = Π*_y_* + Π*_o_*) (b), with continuous (*α_o =_* 0.026) and dashed (*α_o =_* 0.009) curves. At *t =* 120 days, Ω and Π approached asymptotes (or plateaus).

From Figure 4, the first estimation method for additional mortality rates provided around 2.5-times more than the second method. Especially, the first method estimated 26.7% against 11.2% of deaths in the elder subpopulation, while for the young subpopulation, 5% against 2%. Hence, the second estimation method is more credible.

For lower *R*_0 =_ 4.097, Figure 5(a) shows the estimated curves of accumulated number of severe CoViD-19 (Ω*_y_*, Ω*_o_*, and Ω = Ω*_y_* +Ω*_o_*), from equation (13). At *t =* 160 days, Ω approached an asymptote (or plateau), which can be understood as the time when the first wave of epidemics ends. The curves Ω*_y_*, Ω*_o_*, and Ω reach values at *t =* 200, respectively, 1.484 × 10^6^,0.422 × 10^6^, and 1.906 × 10^6^. Figure 5(b) shows the estimated curves of the accumulated number of CoViD-19 deaths (Π*_y_*, Π*_o_*, and Π = Π*_y_* + Π*_o_*), from equation (14). The values of Π*_y_*, Π*_o_*, and Π are at *t =* 200, for the first method of estimation, respectively, 6.594×10^5^ (44.4%), 3.583×10^5^ (84.9%) and 1.018 × 10^6^ (53.41%), and for the second method of estimation, respectively, 2.62 × 10^4^ (1.76%), 4.718 × 10^4^ (11.18%) and 7.338 × 10^4^ (3.85%). The percentage between parentheses is the ratio Π*/*Ω.

From Figure 5, the first estimation method for additional mortality rates provided much higher values than the second method, for instance, 25-times more among young subpopulation. Especially, the first method estimated 85% against 11.2% of deaths in the elder subpopulation. Hence, the second estimation method is more credible. Notice that, from Figures 4 and 5, at the end of the first epidemic wave, we have quite the same number of accumulated CoViD-19 cases for *R*_0 =_ 6.828 and 4.097 (difference at most 1.8%) despite big differences in accumulated deaths.

**Figure 5:**
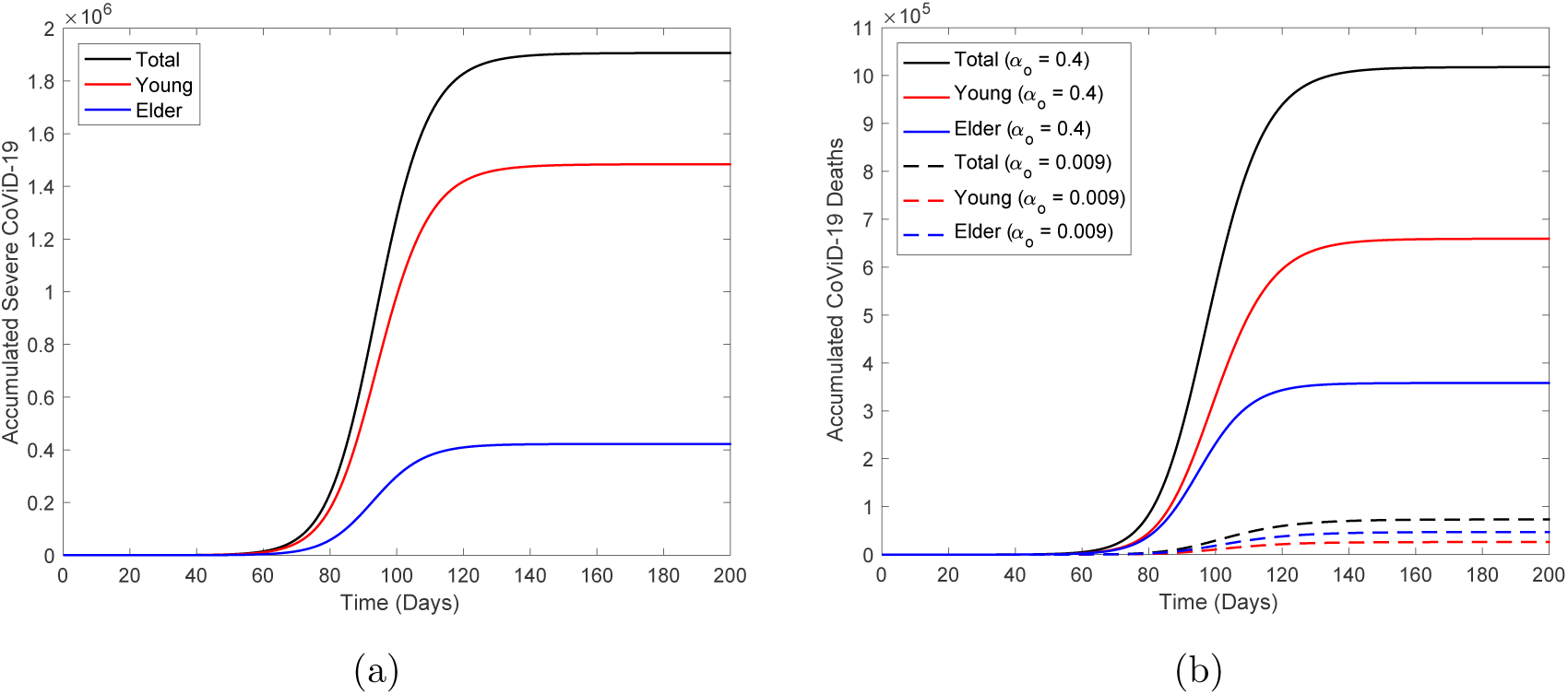
For *R*_0 =_ 4.097, the estimated curves of accumulated number of severe CoViD-19 (Ω*_y_*, Ω*_o_* and Ω = Ω*_y_* +Ω*_o_*) (a), from top to bottom for total, young and elder persons, and accumulated number of CoViD-19 deaths (Π*_y_*, Π*_o_* and Π = Π*_y_* +Π*_o_*) (b), with continuous (*α_o =_* 0.4) and dashed (*α_o =_* 0.009) curves. At *t =* 160 days, Ω and Π approached plateaus.

At *t =* 0, the numbers of susceptible persons, *S_y_*, *S_o_* and *S = S_y_* + *S_o_* are, respectively, 3.77762 × 10^7^,0.68238 × 10^7^ and 4.46 × 10^7^, which diminish due to infection at lower values at *t =* 200 (figure not shown, see Figure 8(a) below to observe sigmoid shape). For higher *R*_0 =_ 6.828, the numbers of susceptible persons left behind are 2.346 × 10^5^ (0.62%), 0.294 × 10^4^ (0.043%) and 2.376 × 10^5^ (0.53%), for young, elder and total persons, respectively. For lower *R*_0 =_ 4.097, the numbers of susceptible persons left behind are 8.823 × 10^5^ (2.34%), 6.957 × 10^4^ (1.02%) and 9.518×10^5^ (2.13%), for young, elder and total persons, respectively. The percentage between parentheses is the ratio *S*(200)*/S*(0). For young and total persons, there are 4-times more susceptible persons available to be infected in the second wave when *R*_0_ decreases, while for elder persons, 24-times. Hence, the second wave will infect much more elder persons.

For higher *R*_0 =_ 6.828, the numbers of immune persons (*I_y_*, *I_o_*, and *I*) increase from zero to, respectively, 3.755 × 10^7^ (99.4%), 0.674 × 10^7^ (98.8%) and 4.429 × 10^7^ (99.3%), for young, elder and total persons at *t =* 200 (figure not shown, see Figure 8(b) below). For lower *R*_0 =_ 4.097, the numbers of immune persons at *t =* 200 are 3.689 × 10^7^ (97.7%), 0.668 × 107 (97.9%) and 4.357 × 10^7^ (97.7%), for young, elder and total persons, respectively. The percentage between parentheses is the ratio *I/S*(0). The value of *R*_0_ does not matter in the number of immune persons.

Remembering that human population is varying due to the additional mortality (fatality) of severe CoViD-19, we have, at *t =* 0, *N*_0_*_y =_* 3.780 × 10_7,_ *N*_0_*_o =_* 0.680 × 10_7_ and *N*_0 =_ *N*_0_*_y_* + *N*_0_*_o =_* 4.460 × 10_7_, and at *t =* 200, *N_y =_* 3.774 × 10_7_ (0.16%), *N_o =_* 0.667 × 10_7_ (1.91%) and *N =* 4.441 × 10_7_ (0.43%) for the first estimation method, and *N_y =_* 3.779 × 10_7_ (0.026%), *N_o =_* 0.674 × 10_7_ (0.88%) and *N =* 4.453 × 10_7_ (0.16%) for the second estimation method. The percentage of deaths is the ratio (*N*_0_*_j_* − *N_j_*) /*N*_0_*_j_*.

#### 3.1.3 Estimation of the proportion of isolated persons

From Figures 4 and 5, as R_0_ decreases, the estimation of the additional mortality rates based on a few initially collected data becomes more inaccurate, providing so much deaths in the population. Hence, we choose *β_y =_* 0.75 and *β_o =_* 0.8775 (higher *R*_0 =_ 6.828) and lower estimates for the mortality rates, *α_y =_* 0.0018 and *α_o =_* 0.009 (all in *days*^−1^), to estimate isolation (described by proportions *k_y_* and *k_o_*) of susceptible persons as control mechanism. Isolation was introduced at *t =* 27 (March 24), and we will estimate taking into account the confirmed cases from *t =* 27 to 55 (April 21), hence *n =* 29 observations.

Here, we estimate the control variables by varying *k_y_* and *k_o_*. The system of equations (2), (3), and (4), with boundary conditions given by equations (8) and (9), is evaluated, and we calculate the sum of square differences

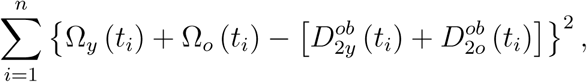

where *t*_1 =_ 27 and *t*_29 =_ 55. We assume *k = k_y =_ k_o_* and varied *k =* 0, 0.4, 0.6, 0.7, and 0.8, from which the better estimated value is *k =* 0.5 [1]. In this section, the isolation was initiated at *t =* 27, and release is not initiated.

Figure 6(a) shows the estimated curve of *D*_2 =_ *D*_2_*_y_* + *D*_2_*_o_* and the observed data, plus four curves with lower and higher values of *k* than that estimated *k =* 0, 0.4, 0.6, 0.7 and 0.8. Figure 6(b) shows the curves of *D*_2_ for 6 different values of *k*, extended from *t =* 0 until 250. For *k =* 0, 0.4, 0.5, 0.6, 0.7 and 0.8, the peaks are, respectively, 6.889 × 10^5^, 3.117 × 10^5^, 2.25 × 10^5^, 1.423 × 10^5^,0.68 × 10^5^ and 0.137 × 10^5^, which decrease as *k* increases, and displace to right, to *t =* 68, 82, 88, 99, 118 and 165.

**Figure 6:**
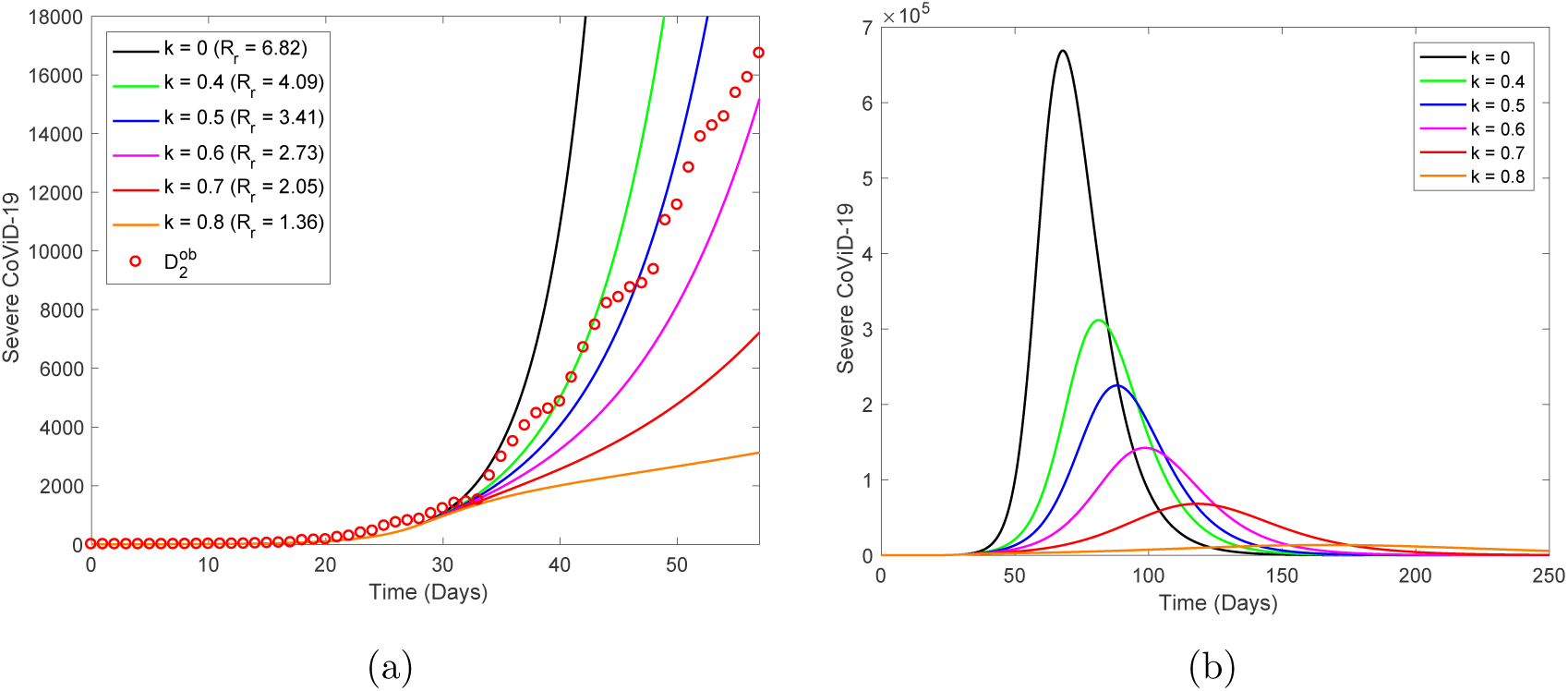
The estimated curve (for *k =* 0.5) of *D*_2_ and observed data, plus five curves with lower and higher values of *k* than that estimated, *k =* 0, 0.4, 0.6, 0.7 and 0.8 (a), and curves of *D*_2_ extended from *t =* 0 until 250 (b), from top to bottom as *k* increases.

Figure 7 shows the curves of Ω*_y_*, Ω*_o_*, and Ω = Ω*_y_* + Ω*_o_* (a) and Π*_y_*, Π*_o_*, and Π = Π*_y_* +Π*_o_* (b) without (*k =* 0) and with isolation (*k =* 0.5). For *k =* 0.5, the curves Ω*_y_*, Ω*_o_*, and Ω attain values at *t =* 250, respectively, 7.296×10^5^,2.09×10^5^ and 9.382×10^5^, and Π*_y_*, Π*_o_* and Π attain, at *t =* 250, respectively, 1.29 × 10_4_ (1.77%), 2.334 × 10_4_ (11.17%) and 3.623 × 10_4_ (3.86%). The percentage between parentheses is the ratio Π*/*Ω.

Figure 8 shows the curves of *S_y_*, *S_o_*, and *S = S_y_* + *S_o_* (a) and *I_y_*, *I_o_*, and *I = I_y_* + *I_o_* (b) without (*k =* 0) and with isolation (*k =* 0.5). The numbers of susceptible persons, *S_y_*, *S_o_*, and *S = S_y_* + *S_o_* are, respectively, at *t =* 250 days, 9.948 × 10^5^ (2.63%), 7.942 × 10_4_ (1.17%) and 1.075 × 10_6_ (2.41%), for *k =* 0.5. The percentage between parentheses is the ratio *S*(250)*/S*(0). The number of immune persons (*I_y_*, *I_o_*, and *I*) increases from zero to, respectively, 1.811 × 10_7_ (47.9%), 0.329 × 10_7_ (48.4%) and 2.141 × 10_7_ (48%), for *k =* 0.5. The percentage between parentheses is the ratio *I*/*S*(0), and *S*(0) can be found in the foregoing section.

From foregoing section, we transport values for *k =* 0: for Ω*_y_*, Ω*_o_*, and Ω = Ω*_y_* + Ω*_o_*, 1.511 × 10_6_,0.426 × 10_6_ and 1.937 × 10_6_; for Π*_y_*, Π*_o_* and Π, 2.67 × 10_4_ (1.77%), 4.767 × 10_4_ (11.19%) and 7.437×10_4_ (3.84%), ratio is Π*/*Ω; *S_y_*, *S_o_*, and *S = S_y_* +*S_o_* (left behind at *t =* 200), 2.346 × 10^5^ (0.62%), 0.294 × 10_4_ (0.043%) and 2.376 × 10^5^ (0.53%), ratio is *S*(200)*/S*(0); and for *I_y_*, *I_o_*, and *I*,3.755 × 10_7_ (99.4%), 0.674 × 10_7_ (98.8%) and 4.429 × 10_7_ (99.3%), ratio is *I/S*(0). At *t =* 0, for *S_y_*, *S_o_*, and *S*, we have 3.77762 × 10_7_, 0.68238 × 10_7_ and 4.46 × 10_7._ From Figures 6–8, the peak of severe CoViD-19 cases decreases from 688.9 thousand to 225 thousand and is delayed in 20 days, from 68 (May 4) to 88 (May 24).

**Figure 7:**
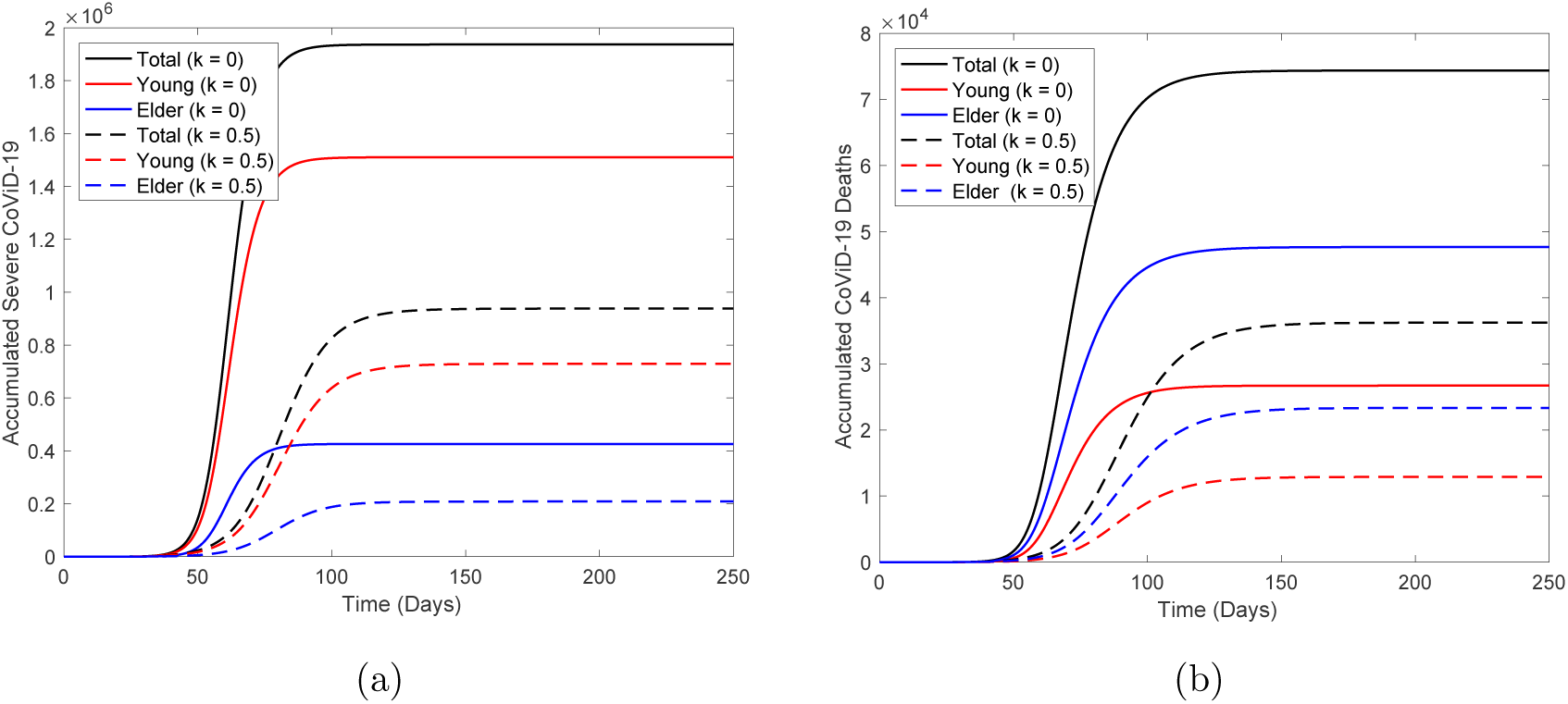
The curves of Ω*_y_*, Ω*_o_* and Ω = Ω*_y_* + Ω*_o_* (a) and Π*_y_*, Π*_o_* and Π = Π*_y_* +Π*_o_* (b) without (*k =* 0, continuous curves) and with isolation proportion (*k =* 0.5, dashed curves), from top to bottom for total, young and elder persons.

As *k* increases to 0.6, 0.7, and 0.8, the peaks decrease to 142.3, 68, and 13.7 (thousand), and are delayed in 31, 50, and 97 (days), occurring at 99 (June 4), 118 (June 23) and 165 (August 9). Severe CoViD-19 cases Ω*_y_*, Ω*_o_*, and Ω = Ω*_y_* + Ω*_o_* decreased to 51%, 49% and 48%, while Π*_y_*, Π*_o_*, and Π, to 48%, 49% and 49%. Susceptible persons *S_y_*, *S_o_*, and *S* left behind increased to 424%, 2, 701% and 453%, while *I_y_*, *I_o_*, and *I*, decreased to 48%, 49% and 48%. During the first wave, isolation decreased the number of severe cases and deaths around 50%, as well as immune persons decreased in 50%. However, susceptible persons left behind increased around 5-times. These pictures show that the next second wave will occur earlier and more intense when compared to *k =* 0.

**Figure 8:**
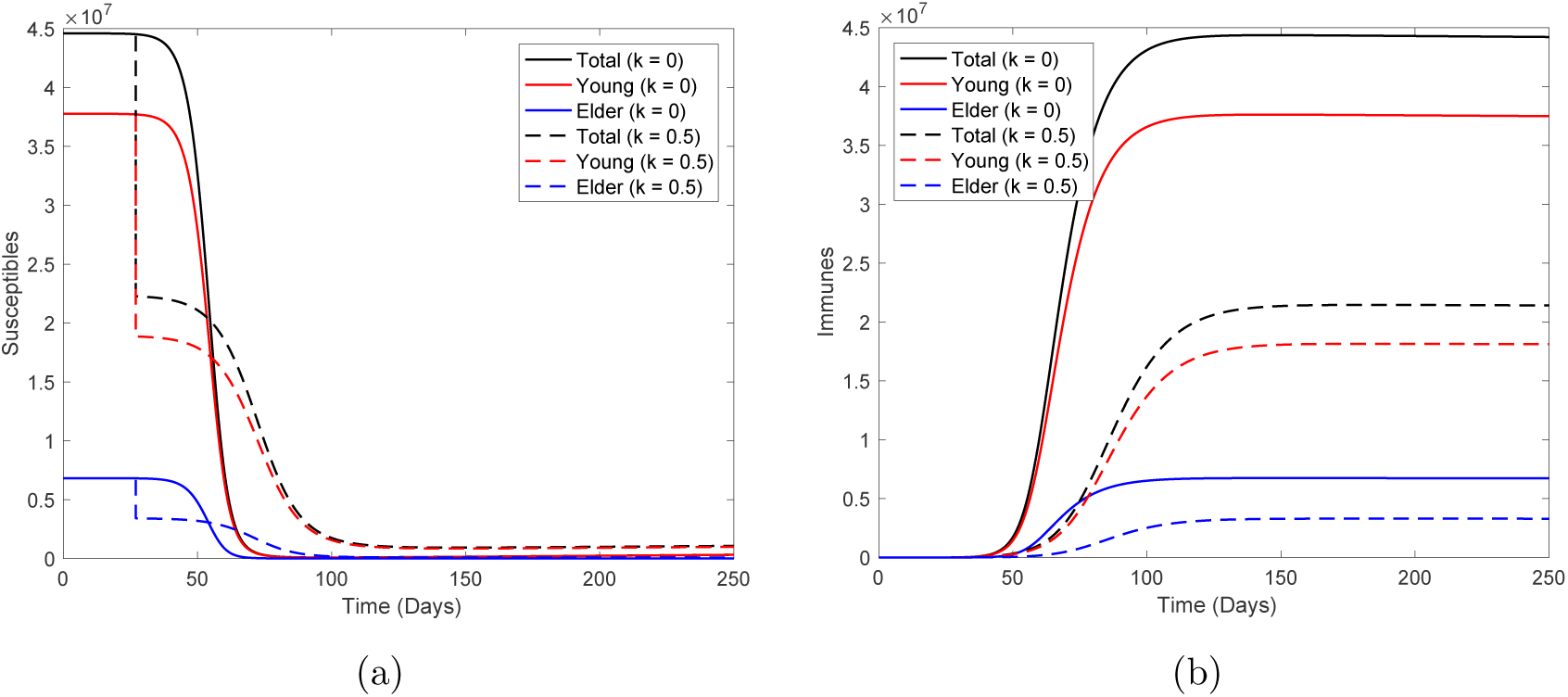
The curves of *S_y_*, *S_o_* and *S = S_y_* + *S_o_* (a) and *I_y_*, *I_o_* and *I = I_y_* + *I_o_* (b) without (*k =* 0, continuous curves) and with isolation proportion (*k =* 0.5, dashed curves), from top to bottom for total, young and elder persons.

Table 3 shows the values of Ω, Π, *S*, and *I* at *t =* 250, for *k =* 0.6, 0.7, and 0.8. Percentages are calculated with respect to *k =* 0. As *k* increases, the numbers of severe CoViD-19 and deaths decrease, which help in the hospital management and care system. However, the remaining susceptible persons increase and less persons are immune, which indicates an anticipation of the second wave. Especially among elder persons, where the number of susceptible persons is so high. Notice that Figure 6 indicates that the number of susceptible persons did not reach the plateau at *t =* 250 when *k =* 0.8.

**Table 3:** The values of Ω, Π, *S* and *I* at *t =* 250, for *k =* 0.6, 0.7 and 0.8. *y*, *o* and Σ stand for, respectively, young, elder and total persons.

| | $k = 0.6$ | | | $k = 0.7$ | | | $k = 0.8$ | | |
| --- | --- | --- | --- | --- | --- | --- | --- | --- | --- |
| | $y$ | $o$ | $\Sigma$ | $y$ | $o$ | $\Sigma$ | $y$ | $o$ | $\Sigma$ |
| $\Omega$ ($10^5$) | 5.582 | 1.615 | 7.197 | 3.719 | 1.099 | 4.818 | 1.432 | 0.442 | 1.874 |
| $\Pi$ | 9866 | 18060 | 27926 | 6568 | 12270 | 18838 | 2451 | 4752 | 7203 |
| $S$ ($10^6$) | 1.507 | 0.153 | 1.66 | 2.387 | 0.298 | 2.685 | 4.242 | 0.654 | 4.896 |
| $I$ ($10^6$) | 13.86 | 2.55 | 16.41 | 9.238 | 1.735 | 10.973 | 3.495 | 0.683 | 4.178 |
| $\Omega$ (%) | 36.94 | 37.89 | 37.15 | 24.61 | 25.79 | 24.87 | 9.48 | 10.37 | 9.67 |
| $\Pi$ (%) | 36.95 | 37.89 | 37.55 | 24.60 | 25.74 | 25.33 | 9.18 | 9.97 | 9.69 |
| $S$ (%) | 477 | 5055 | 520 | 755 | 9845 | 841 | 1342 | 21606 | 1534 |
| $I$ (%) | 36.98 | 37.82 | 37.11 | 24.65 | 25.73 | 24.81 | 9.32 | 10.13 | 9.45 |

Until now, we estimated the model parameters aiming understanding new coronavirus dynamics in the São Paulo State. Next, we evaluate the release of isolated persons, which will occur on May 11.

### 3.2 Epidemiological scenarios considering unique isolation followed by releases

To obtain epidemiological scenarios, we consider the higher *R*_0_ and the second method of estimation for additional mortality rates. Additionally, we choose *k =* 0.5 for the proportion of isolated persons in the São Paulo State. Hence, the fitted values *β_y =_* 0.75, *β_o =_* 0.8775, *α_y =_* 0.0018, *α_o =_* 0.009 (*days*^−1^), and *k =* 0.5 are fixed hereafter, unless explicitly cited.

In this section, considering only one isolation occurred on March 24 (*t =* 27), we study the epidemiological scenarios of release when this isolation will be ended on May 10 (*t =* 74).^1^ We stress the fact that the isolation of susceptible persons taken into account by the model discriminates who has or do not have contact with the circulating new coronavirus. In the absence of mass testing, it is expected that among isolated persons there will be who harbor virus. We simulate the beginning of release occurring at *t =* 72, but also at *t =* 56 (April 22).^2^ We consider 3 strategies of release: all released at *t =* 72 (56) (strategy 1); two releases equally distributed at *t =* 72 (56) and *t =* 79 (63) (strategy 2); and three releases equally distributed at *t =* 72 (56), *t =* 79 (63) and *t =* 86 (70) (strategy 3). We also consider two regimes: equal proportions of release for young and elder persons (regime 1), and different proportions (regime 2).

To obtain the scenarios, we solve numerically the system of equations (2), (3), and (4) with initial conditions (*t =* 0) given by equation (7), the boundary conditions in isolation occurred at *t =* 27 given by equations (8) and (9), and boundary conditions of releases with first one occurring at *t =* 72 given by equations (10) and (11).

Aiming comparison with the absence of isolation, when *k =* 0, the peaks for young and elder persons are 5.024 × 10^5^ and 1.665 × 10^5^, which occur at *t =* 68. When *k =* 0.5, the peaks for young and elder persons are 1.667 × 10^5^ (33%) and 5.835 × 10^4^ (35%), which occur at *t =* 88. Percentage is peak(*k =* 0.5)*/*peak(*k =* 0).

#### 3.2.1 Regime 1 – Equal proportions of releasing young and elder persons

For strategy 1, we have a unique releases with l_1_*_j =_* 1, for strategy 2, two releases with *l*_1_*_j =_* 0.5 and *l*_2_*_j =_* 1, and for strategy 3, three releases with *l*_1_*_j =_* 0.33, *l*_2_*_j =_* 0.5 and *l*_3_*_j =_* 1, *j = y, o*.

Figure 9 shows the curves of *D*_2_ without (*k =* 0) and with (*k =* 0.5) isolation initiated at *t =* 27, but released according to strategy 1. For release beginning at *t =* 56 for young (a) and elder (b) persons, the peaks are, respectively, 4.957 × 10^5^ (99%) and 4.607 × 10^5^ (92%), with peak occurring at *t =* 82; and for release at *t =* 72, 1.641 × 10^5^ (99%) and 1.529 × 10^5^ (92%), with peak occurring at *t =* 92. The release in comparison with only isolation, peaks are anticipated in 6 and delayed at 4 days, respectively, for young and elder persons.

**Figure 9:**
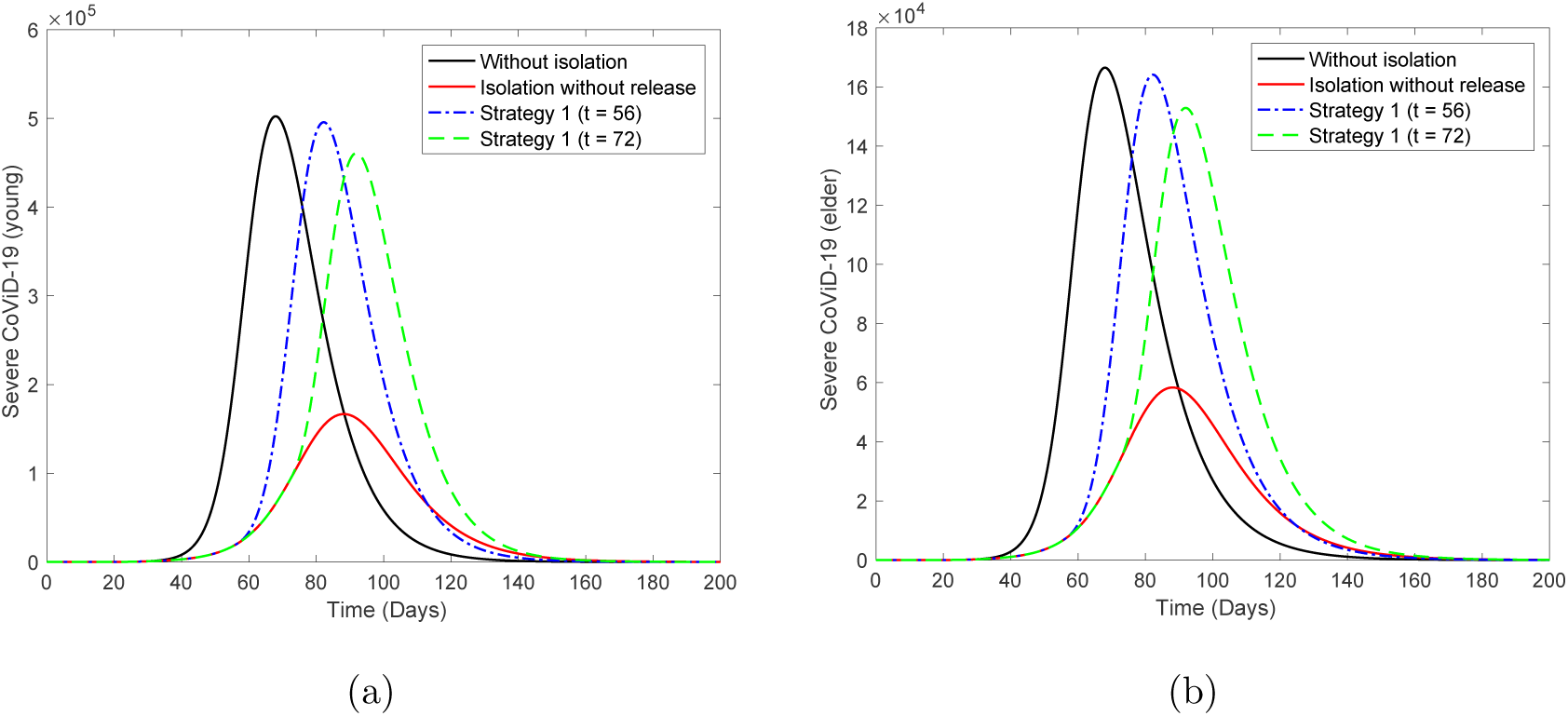
The curves of *D*_2_ without isolation (*k =* 0), with isolation (*k =* 0.5) initiated at *t =* 27, but releasing according to strategy 1 beginning at *t =* 56 (dot and dashed) and 72 (dashed) for young (a) and elder (b) persons. Equal releasing of young and elder persons.

Figure 10 shows the curves of *D*_2_ without isolation (*k =* 0), with isolation (*k =* 0.5) initiated at *t =* 27, but released according to strategy 2. For release beginning at *t =* 56 for young (a) and elder (b) persons, the peaks are, respectively, 4.904 × 10^5^ (98%) and 4.373 × 10^5^ (87%), with peak occurring at *t =* 84; and for release at *t =* 72, 1.624 × 10^5^ (98%) and 1.455 × 10^5^ (87%), with peak occurring at *t =* 94. The release in comparison with only isolation, peaks are anticipated at 4 and delayed in 6 days, respectively, for young and elder persons.

**Figure 10:**
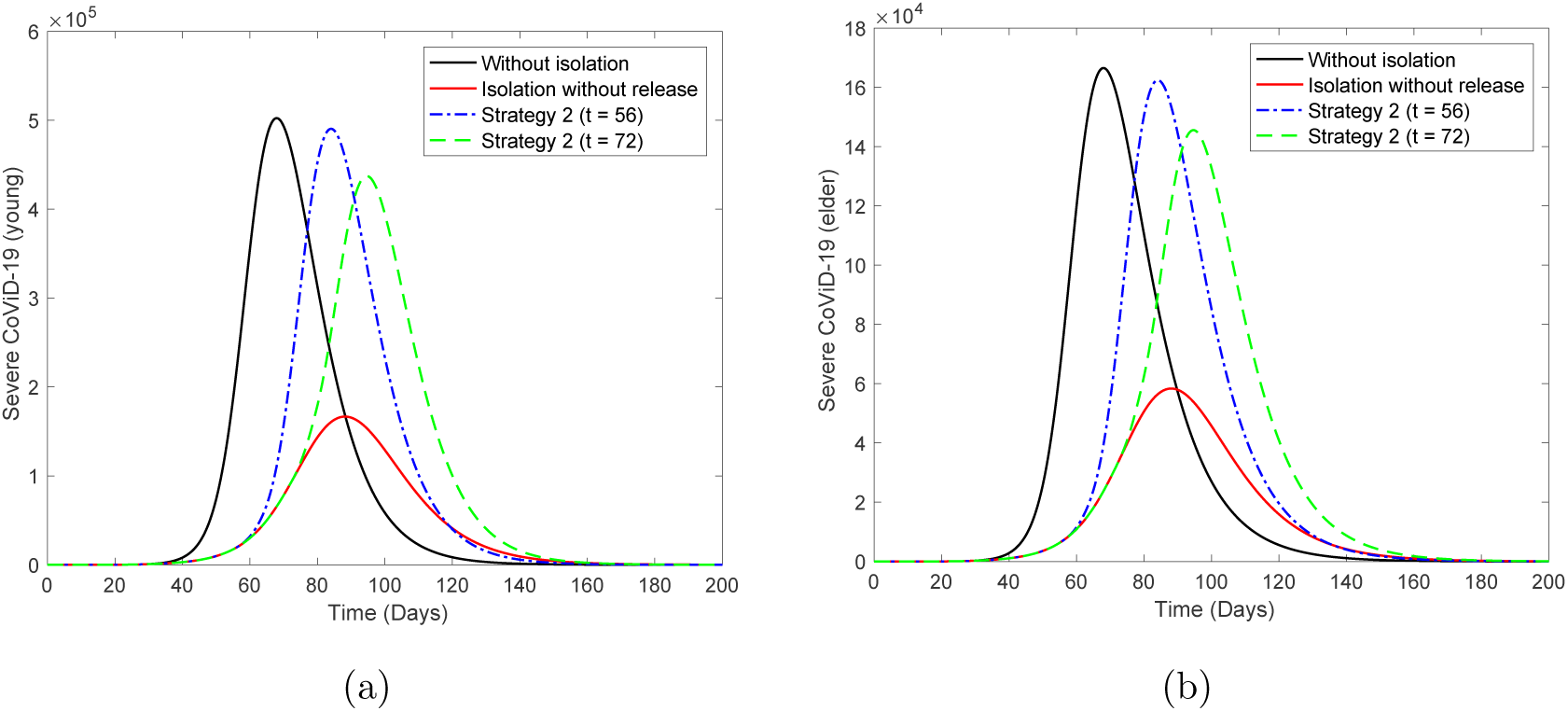
The curves of *D*_2_ without isolation (*k* = 0), with isolation (*k* = 0.5) initiated at *t* = 27, but releasing according to strategy 2 beginning at *t* = 56 (dot and dashed) and 72 (dashed) for young (a) and elder (b) persons. Equal releasing of young and elder persons

Figure 11 shows the curves of *D*_2_ without isolation (*k =* 0), with isolation (*k =* 0.5) initiated at *t =* 27, but release according to strategy 3. For release beginning at *t =* 56 for young (a) and elder (b) persons, the peaks are, respectively, 4.777 × 10^5^ (95%) and 3.97 × 10^5^ (79%), with peak occurring at *t =* 87; and for release at *t =* 72, 1.585 × 10^5^ (95%) and 1.33 × 10^5^ (80%), with peak occurring at *t =* 98. The release in comparison with only isolation, peaks are anticipated at 1 and delayed in 10 days, respectively, for young and elder persons.

**Figure 11:**
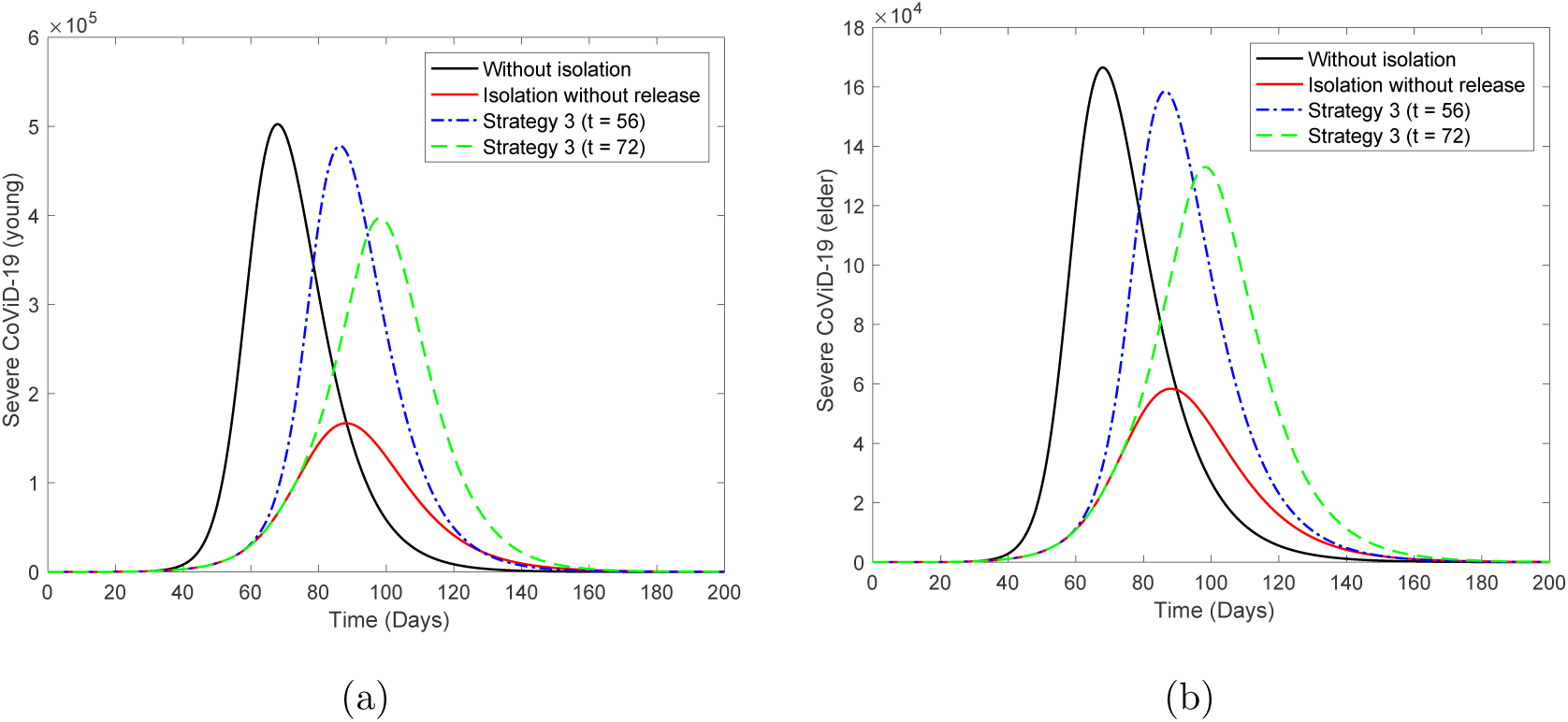
The curves of *D*_2_ without isolation (*k* = 0), with isolation (*k* = 0:5) initiated at *t* = 27, but releasing according to strategy 3 beginning at *t* = 56 (dot and dashed) and 72 (dashed) for young (a) and elder (b) persons. Equal releasing of young and elder persons.

Table 4 shows the values of Ω, Π, *S*, and *I* at *t =* 250, for strategies 1, 2, and 3 for release occurring at *t =* 72. Percentages are calculated with respect to *k =* 0. When young and elder persons are released simultaneously, the numbers of severe CoViD-19 and deaths maintain practically unchanged, as well as immune persons. However, the remaining susceptible persons increase, which indicates that the second wave harm more elder persons. There is very tiny difference among the three strategies.

**Table 4:** Regime 1: the values of Ω, Π, *S an*d *I* at *t =* 250, for strategies 1, 2 and 3 for releasing occurring at *t =* 72. Percentages are calculated with respect to *k =* 0.

|  | Strategy 1 |  |  | Strategy 2 |  |  | Strategy 3 |  |  |
| --- | --- | --- | --- | --- | --- | --- | --- | --- | --- |
| | $y$ | $o$ | $\Sigma$ | $y$ | $o$ | $\Sigma$ | $y$ | $o$ | $\Sigma$ |
| $\Omega$ ($10^6$) | 1.51 | 0.4257 | 1.9357 | 1.509 | 0.4256 | 1.9346 | 1.508 | 0.4254 | 1.9334 |
| $\Pi$ | 26690 | 47610 | 74300 | 26680 | 47600 | 74280 | 26660 | 47570 | 74230 |
| $S$ ($10^5$) | 2.168 | 0.04416 | 2.21216 | 2.288 | 0.057 | 2.345 | 2.615 | 0.09075 | 2.70575 |
| $I$ ($10^7$) | 3.757 | 0.6735 | 4.4305 | 3.756 | 0.6734 | 4.4294 | 3.752 | 0.673 | 4.425 |
| $\Omega$ (%) | 99.93 | 99.88 | 99.92 | 99.87 | 99.86 | 99.87 | 99.80 | 99.81 | 99.80 |
| $\Pi$ (%) | 99.96 | 99.87 | 99.91 | 99.93 | 99.85 | 99.88 | 99.85 | 99.79 | 99.81 |
| $S$ (%) | 92.41 | 150.20 | 93.13 | 97.53 | 193.88 | 98.72 | 111.47 | 308.67 | 113.91 |
| $I$ (%) | 100.05 | 99.90 | 100.03 | 100.03 | 99.88 | 100.00 | 99.92 | 99.82 | 99.91 |

Table 4 presents what happens at the end of the first wave, showing that there is not a significant difference when release is done at *t =* 56 or 72, either for the three strategies. However, during epidemics, release later (*t =* 72) is better, as well as the third strategy.

#### 3.2.2 Regime 2 – Different proportions of releasing young and elder persons

Regime 2 deals with releasing young persons, but maintaining elder persons for a while and releasing all them after young persons. This regime presents quite similar figures observed in regime 1. Hence, we illustrate only one case.

Figure 12 shows the curves of *D*_2_ without isolation (*k =* 0), with isolation (*k =* 0.5) initiated at *t =* 27, but release young persons according to strategy 3 beginning at *t =* 56 (dot and dashed) and 72 (dashed) for young (a) and elder (b) persons. All elder persons are released 21 days after the beginning of releasing young persons, that is, at *t =* 77 and 95. For release beginning at *t =* 56 for young (a) and elder (b) persons, the peaks are, respectively, 4.67 × 10^5^ (93%) and 3.867 × 10^5^ (77%), which occur at *t =* 87 and 98; and release beginning at *t =* 72 for young (a) and elder (b) persons, 1.539 × 10^5^ (92%) and 1.176 × 10^5^ (71%), which occur at *t =* 92 and 109. The release in comparison with only isolation, peaks are anticipated in 1 and delayed in 10 days (*t =* 77), respectively, for young and elder persons, and delayed in 4 and 21 days (*t =* 99) respectively, for young and elder persons. Comparing with Figure 11, the epidemic is slightly mild, but elder persons are infected lately, which could be a desirable effect.

**Figure 12:**
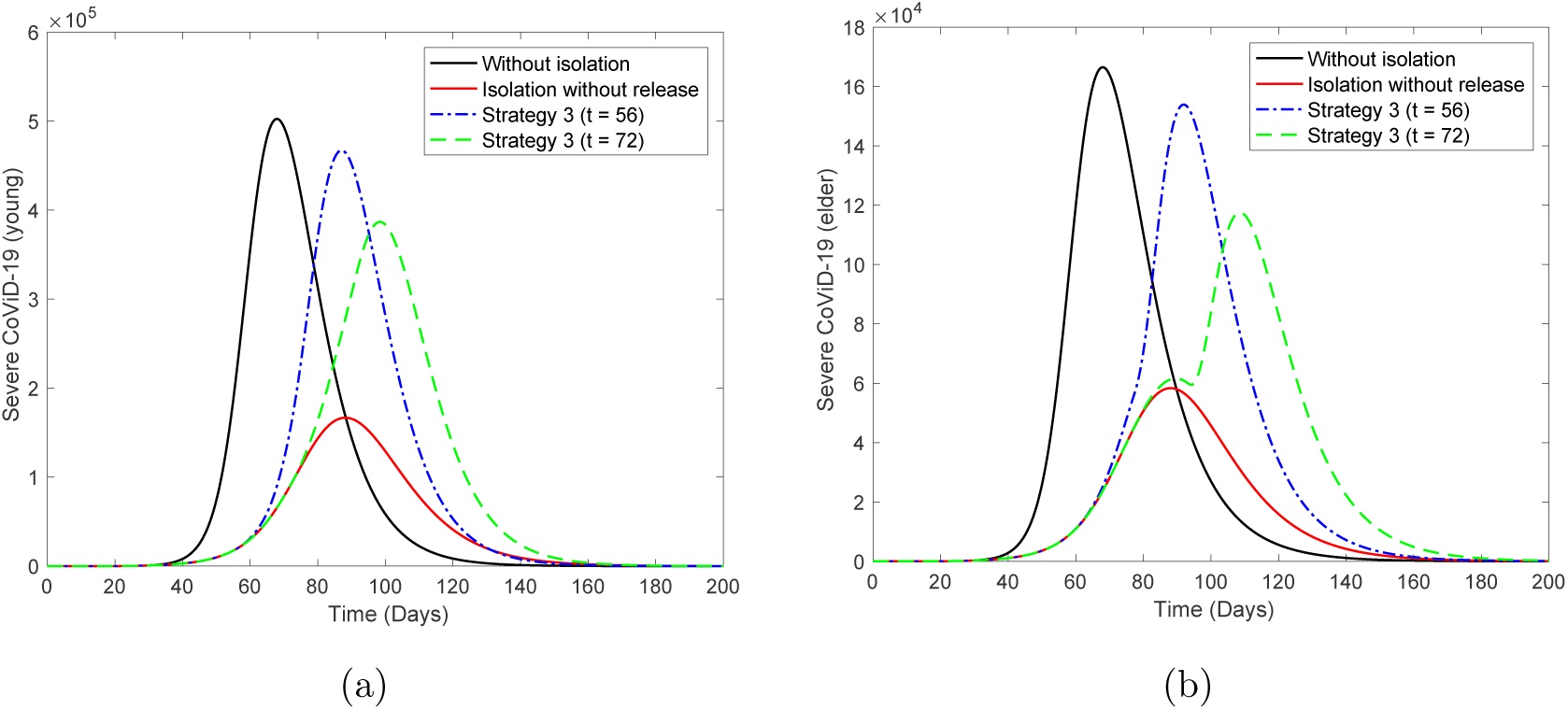
The curves of *D*_2_ without isolation (*k* = 0), with isolation (*k* = 0.5) initiated at *t* = 27, but releasing young persons according to strategy 3 beginning at *t* = 56 (dot and dashed) and 72 (dashed) for young (a) and elder (b) persons. All elder persons are released 21 days after the beginning of release among young persons, that is, at *t* = 77 and 93.

Now, we assess epidemiological scenarios when elder persons are maintained isolated until end of the first wave *t =* 250, but young persons are released according to three strategies, that is, for strategy 1, we have a unique *l*_1_*_y =_* 1, for strategy 2, *l*_1_*_y =_* 0.5 and *l*_2_*_y =_* 1, and for strategy 3, *l*_1_*_y =_* 0.33, *l*_2_*_y =_* 0.5 and *l*_3_*_y =_* 1. Elder persons remain isolated, hence *l_io =_* 0, for all *i*.

Figure 13 shows the curves of *D*_2_ without isolation (*k =* 0), with isolation (*k =* 0.5) initiated at *t =* 27, but release according to strategy 1. For release beginning at *t =* 56 for young (a) and elder (b) persons, the peaks are, respectively, 4.81 × 10^5^ and 4.493 × 10^5^, which occur at *t =* 83 and 92; and for release at *t =* 72 for young (a) and elder (b) persons, 7.714 × 10_4_ and 6.623 × 10_4_, which occur at *t =* 83 and 89. There is minor increasing in epidemic among elder persons (peaks are anticipated) due to the increased number of susceptible young persons.

**Figure 13:**
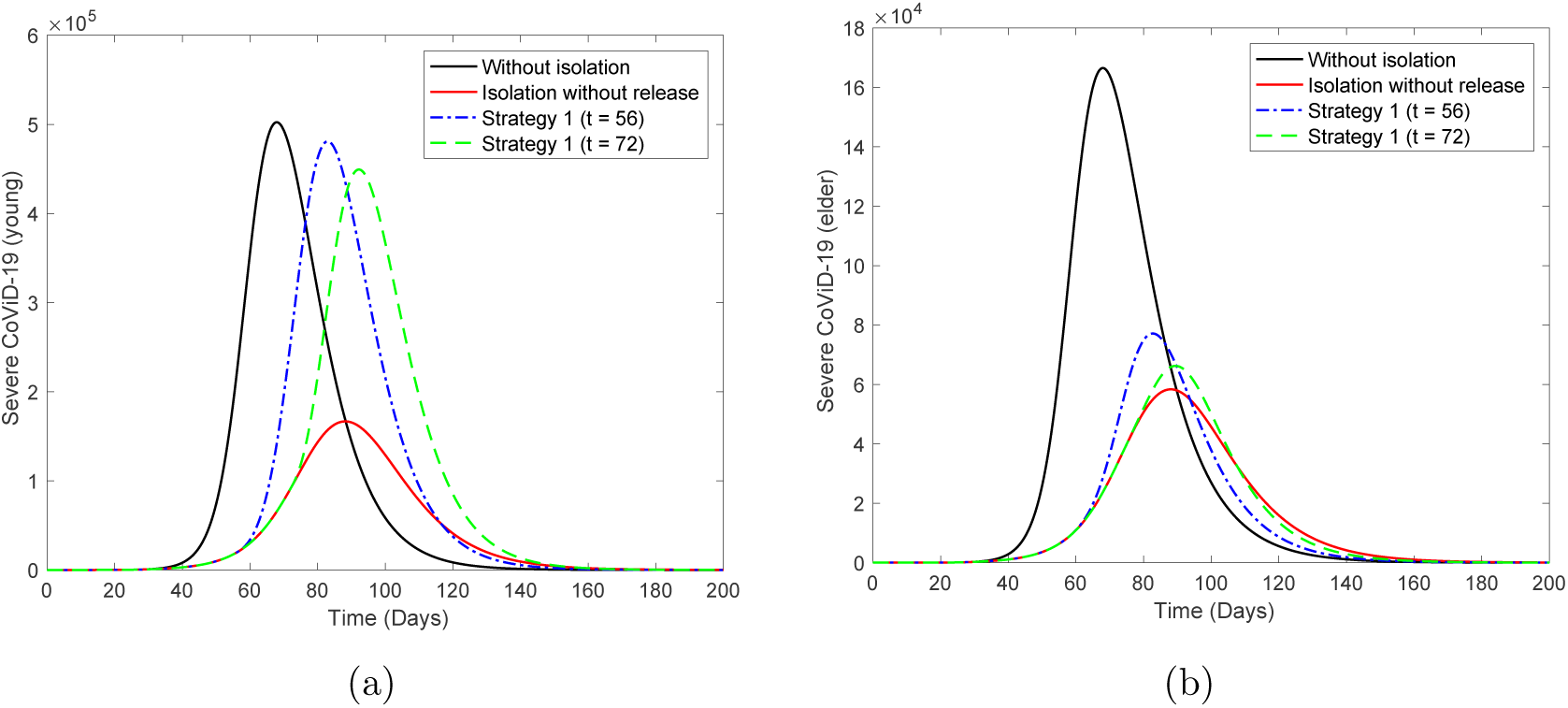
The curves of *D*_2_ without isolation (*k* = 0), with isolation (*k* = 0.5) initiated at *t* = 27, but releasing according to strategy 1 beginning at *t* = 56 (dot and dashed) and 72 (dashed) for young (a) and elder (b) persons. Releasing only young persons.

Figure 14 shows the curves of *D*_2_ without isolation (*k =* 0), with isolation (*k =* 0.5) initiated at *t =* 27, but released according to strategy 2. For release beginning at *t =* 56 for young (a) and elder (b) persons, the peaks are, respectively, 4.764 × 10^5^ and 4.261 × 10^5^, which occur at *t =* 85 and 95; and released at *t =* 72 for young (a) and elder (b) persons, 7.532 × 10^4^ and 6.3514.81 × 10^4^, which occur at *t =* 84 and 90. There is minor increasings in epidemic among elder persons (peaks are anticipated) due to the increased number of susceptible young persons.

**Figure 14:**
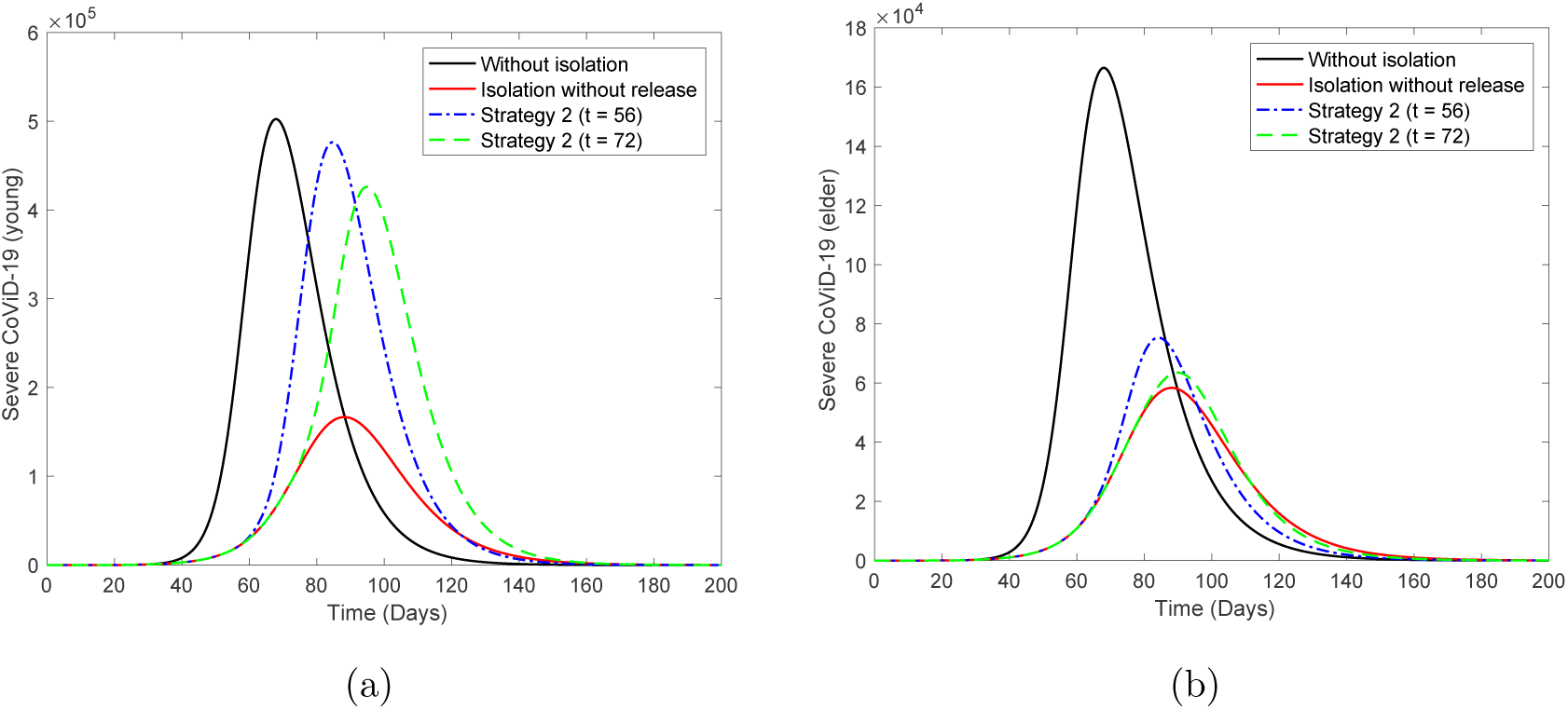
The curves of *D*_2_ without isolation (*k* = 0), with isolation (*k* = 0.5) initiated at *t* = 27, but releasing according to strategy 2 beginning at *t* = 56 (dot and dashed) and 72 (dashed) for young (a) and elder (b) persons. Releasing only young persons.

Figure 15 shows the curves of *D*_2_ without isolation (*k =* 0), with isolation (*k =* 0.5) initiated at *t =* 27, but release according to strategy 3. For release beginning at *t =* 56 for young (a) and elder (b) persons, the peaks are, respectively, 4.652 × 10^5^ and 3.865 × 10^5^, which occur at *t =* 87 and 98; and for release at *t =* 72 for young (a) and elder (b) persons, 7.276 × 10^4^ and 6.158 × 10^4^, which occur at *t =* 86 and 89. There is minor increasings in the epidemic among elder persons (peaks are anticipated) due to the increased number of susceptible young persons released.

**Figure 15:**
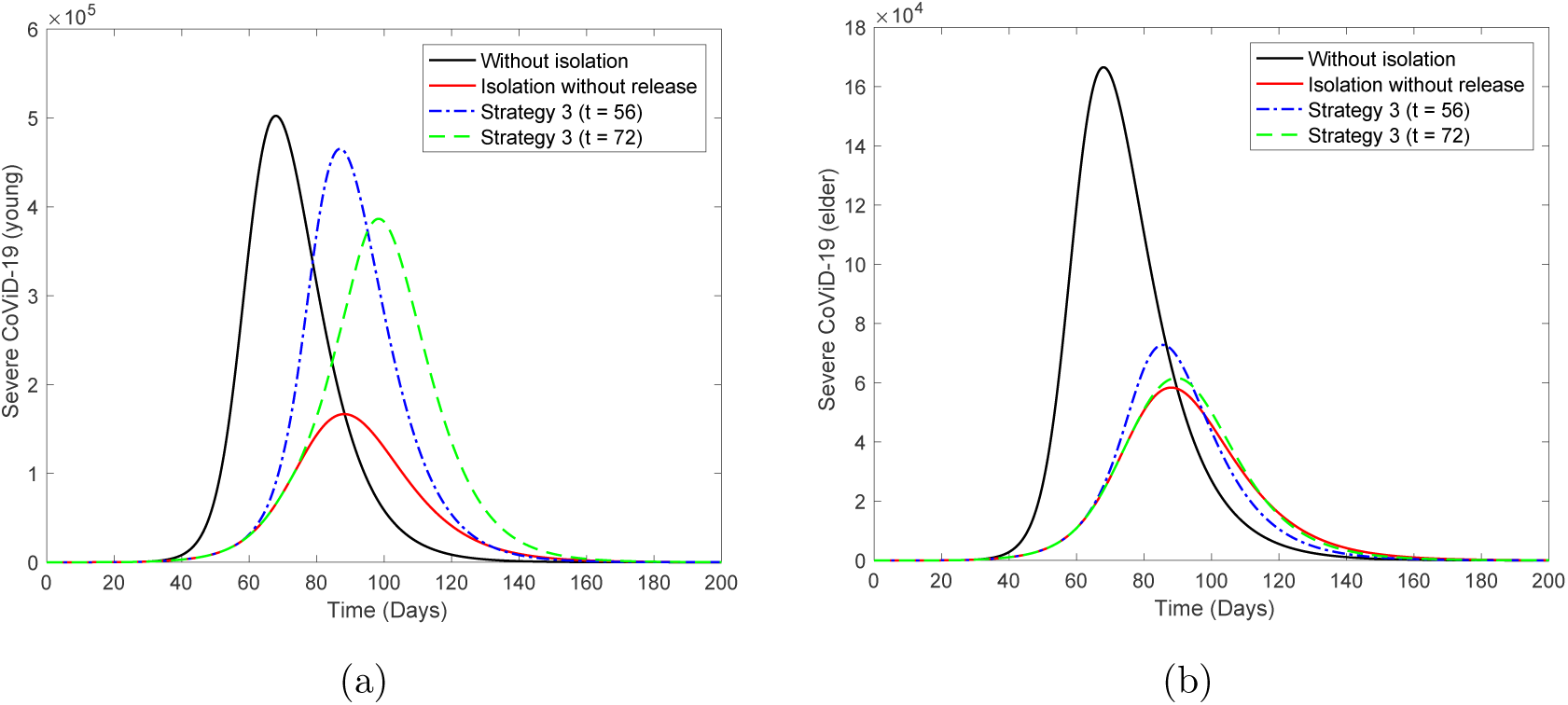
The curves of *D*_2_ without isolation (*k* = 0), with isolation (*k* = 0:5) initiated at *t* = 27, but releasing according to strategy 3 beginning at *t* = 56 (dot and dashed) and 72 (dashed) for young (a) and elder (b) persons. Releasing only young persons.

Table 5 shows the values of Ω, Π, *S*, and *I* at *t =* 250, for strategies 1, 2, and 3 for release occurring at *t =* 72. Percentages are calculated with respect to *k =* 0. When young persons are released while elder persons are maintained isolated, for young classes, the numbers of severe CoViD-19 and deaths maintain practically unchanged, but there are slightly decreasing in immune and increasing in susceptible persons. However, for elder persons there is huge decreasing in the numbers of severe CoViD-19 and deaths, and overall decreasing in severe cases and deaths. There is a tiny difference among the three strategies.

**Table 5:** Regime 2: the values of Ω, Π, *S* and *I* at *t =* 250, for strategies 1, 2 and 3 for releasing occurring at *t =* 72. Percentages are calculated with respect to *k =* 0.

|  | Strategy 1 |  |  | Strategy 2 |  |  | Strategy 3 |  |  |
| --- | --- | --- | --- | --- | --- | --- | --- | --- | --- |
| | $y$ | $o$ | $\Sigma$ | $y$ | $o$ | $\Sigma$ | $y$ | $o$ | $\Sigma$ |
| $\Omega (10^6)$ | 1.508 | 0.2135 | 1.7215 | 1.507 | 0.2135 | 1.7205 | 1.505 | 0.2135 | 1.7185 |
| $\Pi$ | 26660 | 23870 | 50530 | 26640 | 23870 | 50510 | 26600 | 23870 | 50470 |
| $S (10^5)$ | 2.67 | 0.02795 | 2.69795 | 2.898 | 0.02823 | 2.92623 | 3.452 | 0.02887 | 3.48087 |
| $I (10^7)$ | 3.752 | 0.3377 | 4.0897 | 3.75 | 0.3377 | 4.0877 | 3.744 | 0.3377 | 4.0817 |
| $\Omega (\%)$ | 99.80 | 50.09 | 88.87 | 99.74 | 50.09 | 88.81 | 99.60 | 50.09 | 88.71 |
| $\Pi (\%)$ | 99.85 | 50.07 | 67.94 | 99.78 | 50.07 | 67.92 | 99.63 | 50.07 | 67.86 |
| $S (\%)$ | 113.81 | 95.07 | 113.58 | 123.53 | 96.02 | 123.19 | 147.14 | 98.20 | 146.54 |
| $I (\%)$ | 99.92 | 50.09 | 92.33 | 99.87 | 50.09 | 92.29 | 99.71 | 50.09 | 92.15 |

Comparing Tables 4 and 5, regime 2 shows an advantage with respect to regime 1 by decreasing the number of severe cases and deaths, and, additionally, delaying the peak of epidemics among elder persons. Instead of maintaining isolated indefinitely, we release all elder persons at 4 different times. The first (*t =* 90) is just after the peak of epidemics without release, the second (*t =* 120) when occurs inflexion, the third (*t =* 130) and the fourth (*t =* 140) when epidemics are ending.

Figure 16 shows the curves of *D*_2_ for elder persons without isolation (*k =* 0), with isolation (*k =* 0.5) initiated at *t =* 27, but release of young persons according to strategy 3 beginning at *t =* 56 (dot and dashed) and 72 (dashed). Disregarding the beginning of release, all elder persons are released at *t =* 90 (a), 120 (b), 130 (c), and 140 (d).

**Figure 16:**
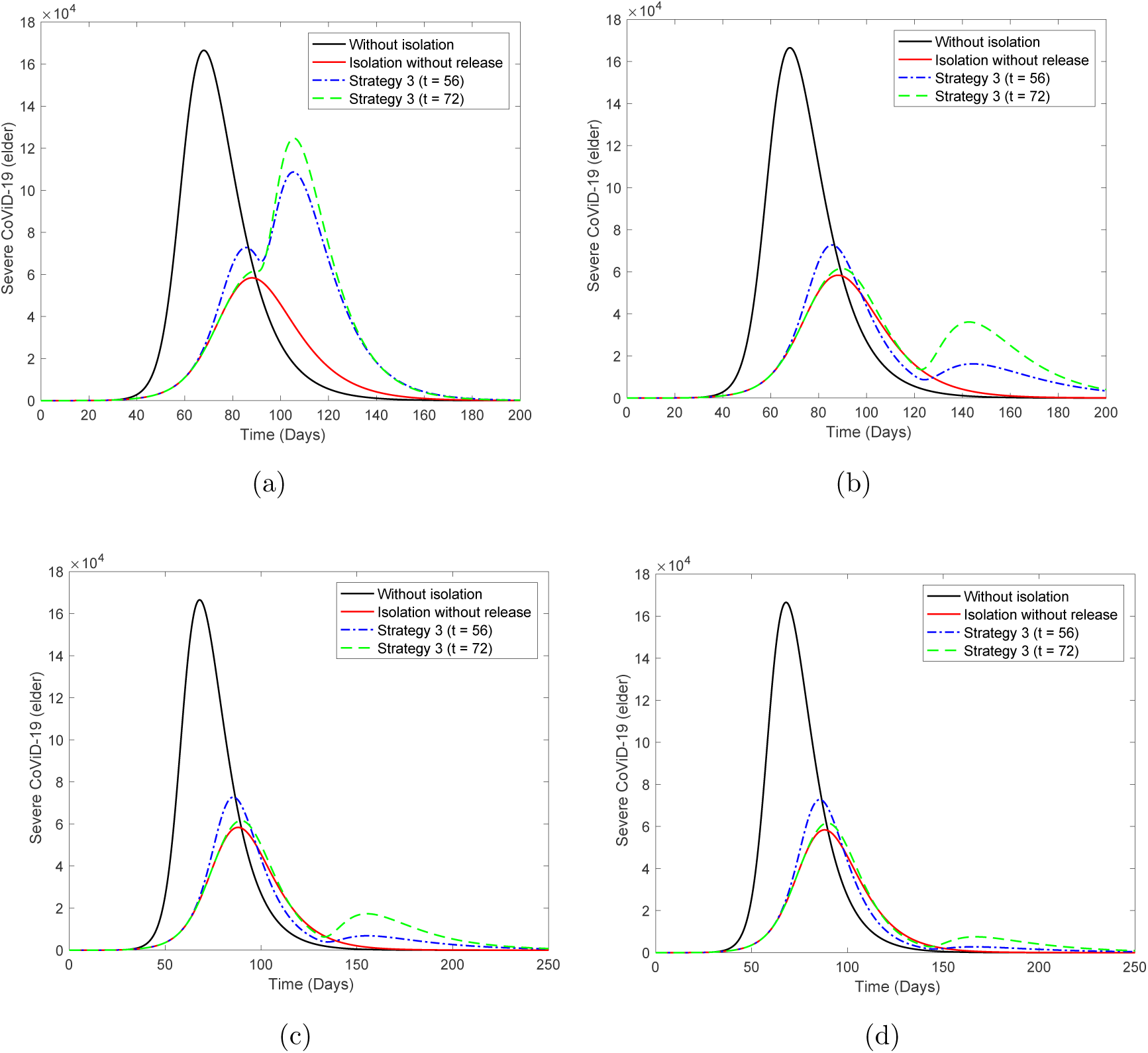
The curves of *D*_2_ for elder persons without isolation (*k* = 0), with isolation (*k* = 0.5) initiated at *t* = 27, but releasing of young persons according to strategy 3 beginning at *t* = 56 (dot and dashed) and 72 (dashed). Disregarding the beginning releasing time, all elder persons are released at *t* = 90 (a), 120 (b), 130 (c) and 140 (d).

As elder persons are released lately, they are protected, and the peak decreases and moves rightwardly with respect to young persons, making more easier the efforts of hospitals to face the pandemic outbreaks. However, the second epidemics wave may occur earlier.

Figure 17 illustrates the pulse release of persons by regime 1 for young persons (a) and regime 2 (b), taking into account strategy 3. Curves without (*k =* 0) and with (*k =* 0.5) isolation are included to observe difference in the dynamics.

**Figure 17:**
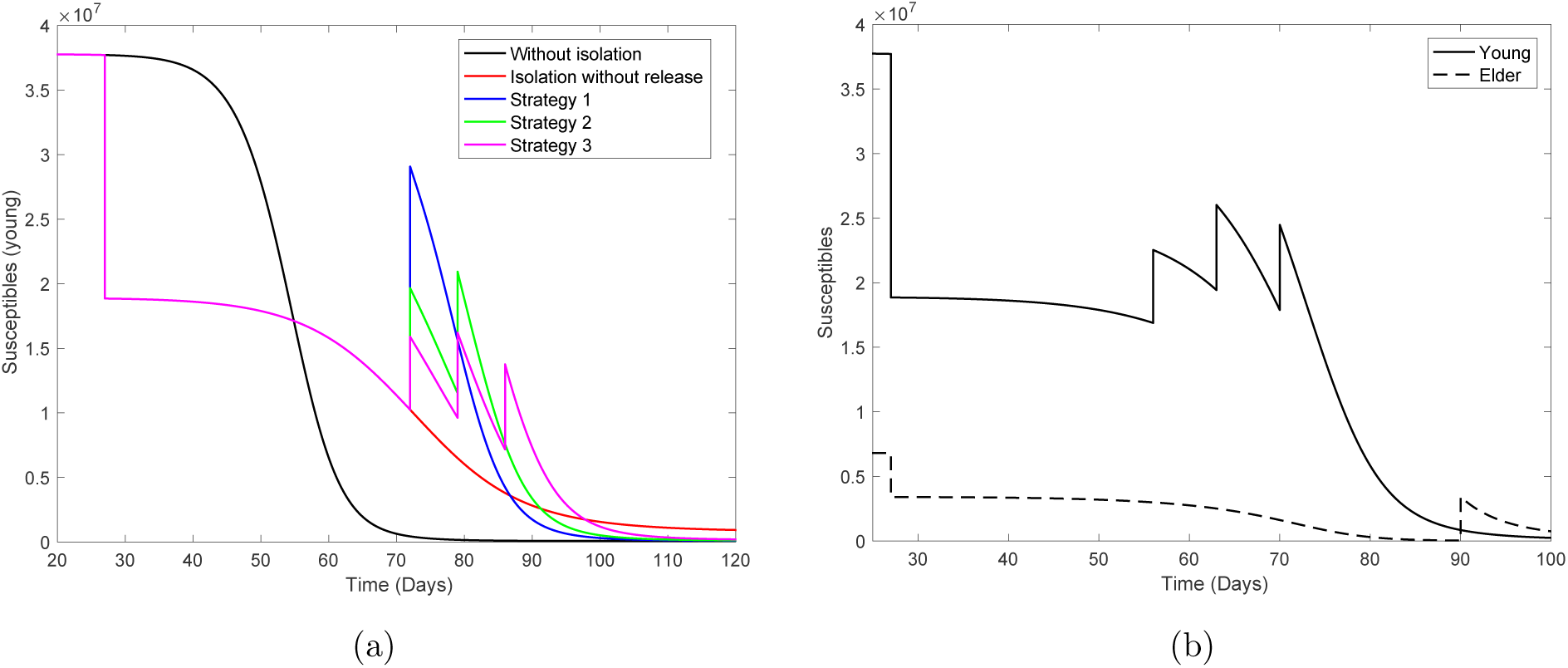
Illustration of pulse release of persons by regime 1 for young persons (a) and regime 2 (b). We show the strategy 3 including curves without (*k* = 0) and with (*k* = 0:5) isolation.

## 4 Discussion

The system of equations (2), (3), and (4) is simulated to provide epidemiological scenarios using parameters estimated from data collected in the São Paulo State, daily released by the Ministry of Health of Brazil. However, these scenarios are more reliable if based on credible values assigned to the model parameters (see discussion in [21]). Here, when we used lower estimated transmission rates, *R*_0 =_ 4.097, to estimate lethality rates against the data, we obtained *α_y =_* 0.08 and *α_o =_* 0.4(*days*^−1^). Using these estimated fatality rates and lower *R*_0_, at the end of the first wave of epidemics, we obtained an incredible 85% of deaths among elder persons.

The substantial difference in the number of deaths relies on the fact that fatality rates are per-capita. In another word, if the number of diseased persons is high, the small fatality rate results in an elevate number of deaths in comparison with the small amount of population. However, the transmission rate is not influenced by the phase of epidemic, due to the fact that this rate multiplies susceptible and infectious individuals.

We assumed that elder persons are under higher risk of being infected by the new coronavirus than young persons, which can be withdrawn if it does not match with actual evidences. Our model did not take into account re-infection (or, maybe relapse), neither if the asymptomatic persons will ultimately manifest symptoms. These unanswered questions do not affect the dynamics during the first wave of epidemics (see [18] for a description of parameters influencing short-term epidemics, and other parameters driving long-term).

Considering that 50% of persons were isolated at the simulation time *t =* 27 (March 27), but they will be released on May 8 (*t =* 72), we draw some epidemiological scenarios. We analyzed three release strategies and concluded that there is little difference among them, but the best scenario was obtained if we release young persons maintaining elder persons isolated. In this case, the number of deaths due to CoViD-19 was reduced by half.

In all scenarios of release with *k =* 0.5, we observed the collapse of the health care system. However, if we increase the proportion of isolated persons above 80%, then the health care system can treat well all patients during the first wave. Let us evaluate the occupancy of beds in hospitals and ICU, using equations (16) and (17). We let *h*_1 =_ *h*_1_*_y =_ h*_1_*_o =_* 1, *h = h_y =_ h_o =_* 0.3, ζ_1 =_ 1*/*14 (2 weeks of hospital care), and ζ_2 =_ 1*/*21 (3 weeks of ICU care) [3].

Figure 18 shows the number of beds occupied in hospitals (a) and ICU (b) for *k =* 0.5 and *k =* 0.7 (dashed curves), and beds occupied in hospitals (c) and ICU (d) for *k =* 0.8. For *k =* 0.5, the peaks of occupied beds in hospitals and ICU (between parentheses) are 1.488 × 10^5^ (8.226 × 10^4^), 4.86 × 10^4^ (2.216 × 10^4^) and 1.895 × 10^5^ (1.043 × 10^5^), occurring at *t =* 90 (92), 88 (90) and 90 (92), respectively for young, elder and total persons. For *k =* 0.7, the peaks of occupied beds in hospitals and ICU (between parentheses) are, respectively, for young, elder, and total persons, 4.646×10^4^ (2.728×10^4^), 1.29×10^4^ (0.738×10^4^) and 5.931×10^4^ (3.462×10^4^), occurring at *t =* 121 (125), 118 (121) and 120 (124). For *k =* 0.8, the peaks of occupied beds in hospitals and ICU (between parentheses) are 9, 495 (5, 879), 2, 674 (1, 597) and 12, 160 (7, 471), occurring at *t =* 170 (176), 165 (169) and 169 (174), respectively for young, elder and total persons.

**Figure 18:**
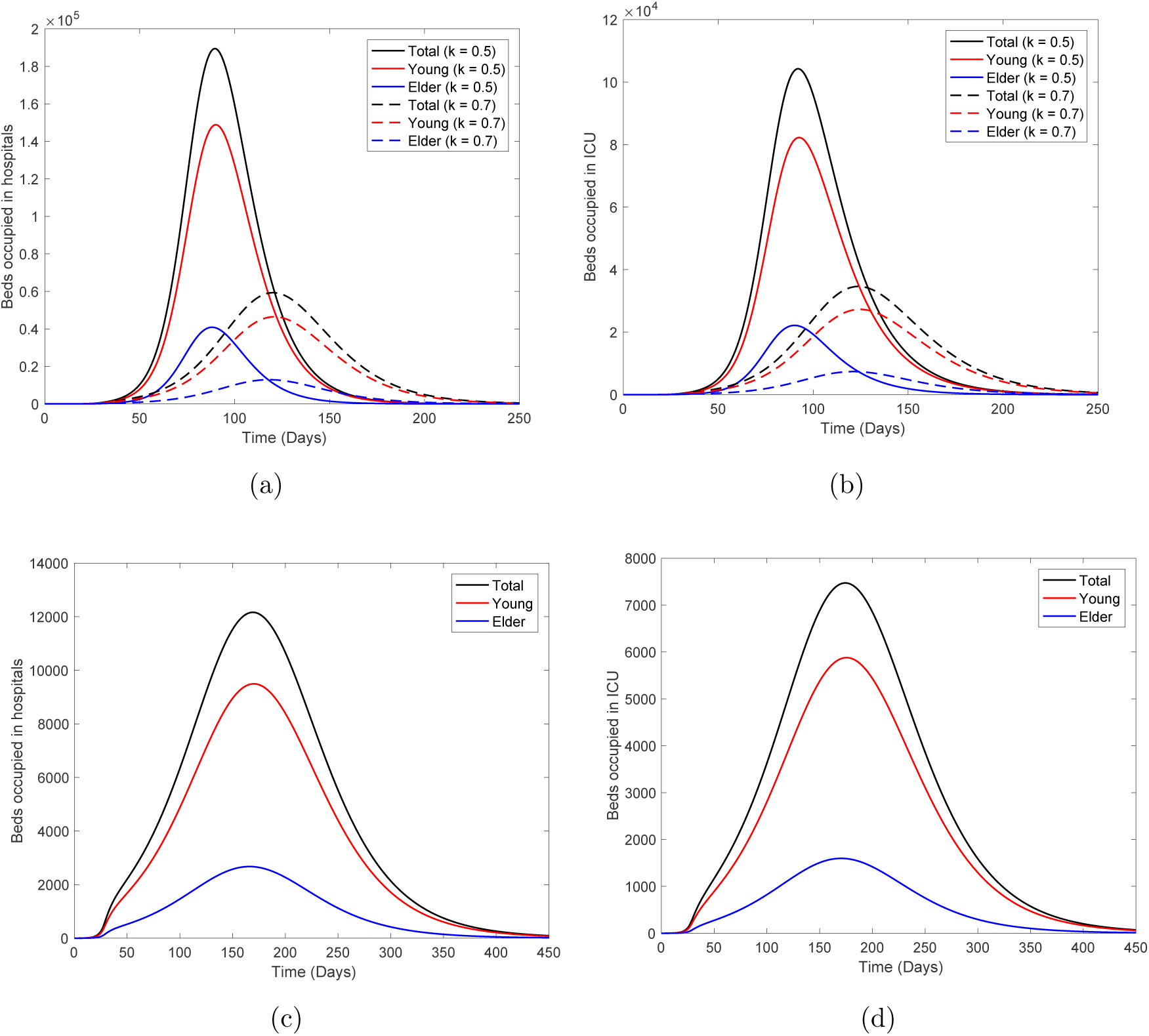
The number of beds occupied in hospitals (a) and ICU (b) for *k* = 0.5 and *k* = 0.7 (dashed curves), and beds occupied in hospitals (c) and ICU (d) for *k* = 0:8.

For *k =* 0.5, the peaks of occupied beds by all persons in hospital and ICU are 189, 500 and 104, 300, occurring at *t =* 90 (May 26) and 92 (May 28). For *k =* 0.7, the peaks of occupied beds by all persons in hospital and ICU are 59, 310 and 34, 620, occurring at *t =* 120 (June 25) and 124 (June 29). For *k =* 0.8, the peaks of occupied beds by all persons in hospital and ICU are 12, 160 and 7, 471, occurring at *t =* 169 (August 13) and 174 (August 18). In this case, From Figure 18, the peak of severe CoViD-19 cases is 13, 700 occurring at *t =* 165 (August 9).

As we have pointed above, when isolation is done lately, among isolated persons, there will be persons harboring virus. Just after the beginning of isolation (*t = τ^is+^*), the number of persons in each class, for *k =* 0.5, is

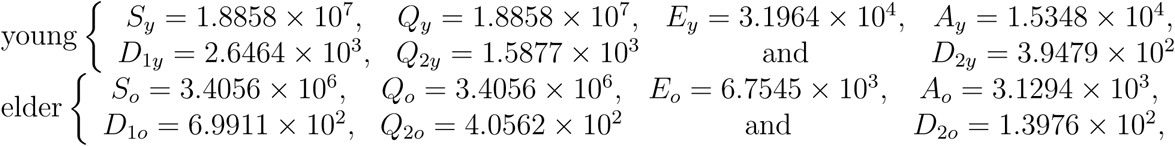

with *Q*_1_*_y =_ Q*_1_*_o =_* 0 and *I =* 8.8376 × 10^3^. To estimate the circulation of virus among isolated persons, we simulate the system of equations (2), (3), and (4) taking as initial conditions the number of persons in each class, at *t = τ^is^*^+^: *S_y =_ Q_y =_* 1.8858 × 10^7^, *S_o =_ Q_o =_* 3.4056 × 10^6^, *Q_y =_ Q_o =_* 0, and for all other variables, half of the corresponding values at *t = τ^is^*^+^. Figure 19 shows the curves of *D*_2_*_y_*, *D*_2_*_o_*, and *D*_2 =_ *D*_2_*_y_* + *D*_2_*_o_* for 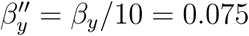, with *R*_0 =_ 1.37 (a), and 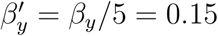, with *R*_0 =_ 0.68 (b), from *t =* 27 to 74, during the period of isolation. At *t =* 72, the numbers of severe CoViD-19 cases are, 1, 597, 635 and 2, 231, respectively, for young, elder, and total persons for *R*_0 =_ 1.37; and 294, 129 and 423, for *R*_0 =_ 0.68. For *k =* 0.5 and *R*_0 =_ 1.37, this 0.8% additional CoViD-19 cases compared to the peak is negligible, however for *k =* 0.8, the 18% additional CoViD-19 cases are not negligible. Nevertheless, when these isolated infectious individuals are released, they contribute to increase the velocity of propagation.

**Figure 19:**
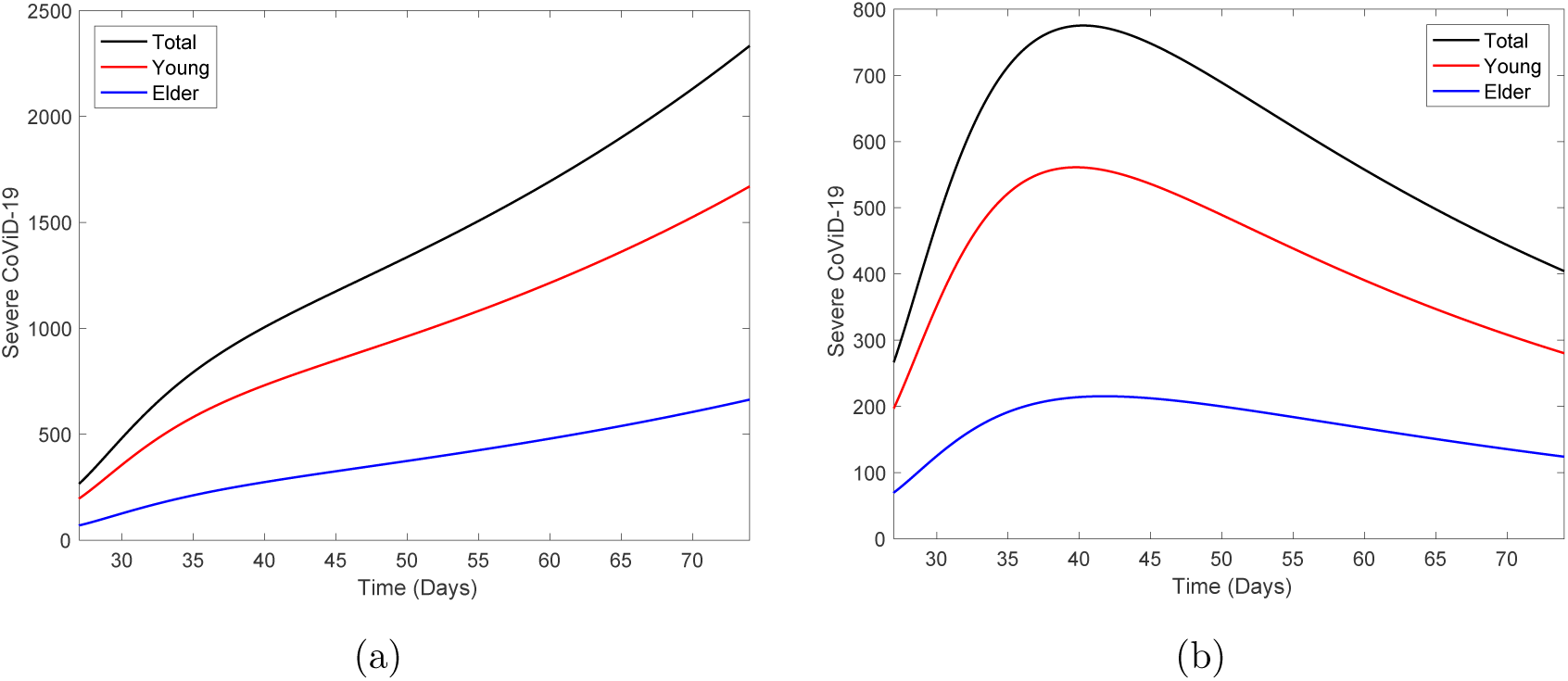
The curves of *D*_2_*_y_*, *D*_2_*_o_* and *D*_2_ = *D*_2_*_y_* + *D*_2_*_o_* for 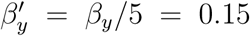 (a) and 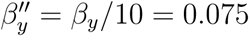 (b) from *t =* 27 to 74, during the period of isolation.

The virus should be circulating among isolated persons through restricted contact occurring in the household and/or neighborhood. If symptomatic persons are transferred to hospital, then removed from isolation, it is expected that asymptomatic persons are predominantly spreading the new coronavirus. If we can assume that they are releasing a low amount of virus, it is expected that more asymptomatic cases will arise than severe CoViD-19. Increasing immune persons, they decrease susceptible persons, hence the effective reproduction number decreases, which is a herd immunity phenomenon.

Let us discriminate the circulation of the new coronavirus in a community. Figure 20 shows all persons harboring this virus (*E_j_*, *A_j_*, *D*_1_*_j_*, *Q*_2_*_j_*, and *D*_2_*_j_*), *j = y, o*, for young (a) and elder (b) persons corresponding to Figure 6(b) with *k =* 0.5. There is little difference: exposed (*E*) and pre-diseased (*D*_1_) persons are relatively higher among young persons.

**Figure 20:**
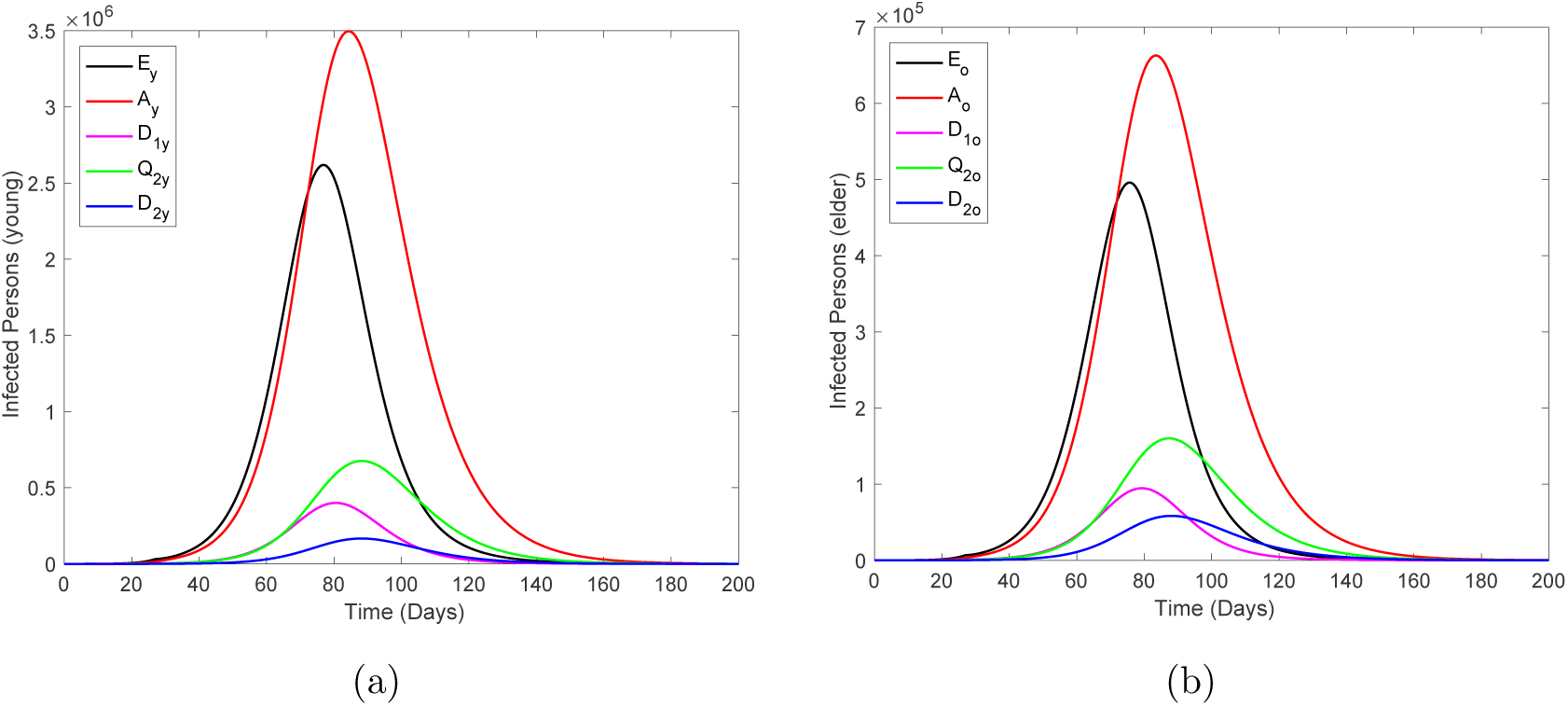
The curves of all persons harboring this virus (*E_j_*, *A_j_*, *D*_1_*_j_*, *Q*_2_*_j_* and *D*_2_*_j_*), *j* = *y*; *o*, for young (a) and elder (b) persons.

In Figure 21 we show the ratio hidden:apparent based on Figure 20. Who harbor the new coronavirus as exposed or who do not manifest symptoms are classified in the hidden category, and in the apparent category, we include who manifests symptoms. Hence, the ratio is calculated as (*E* + *A* + *D*_1_):(*Q*_2_ + *D*_2_). At *t =* 0, the ratio is 10 : 1 for young and elder persons due to initial conditions.

**Figure 21:**
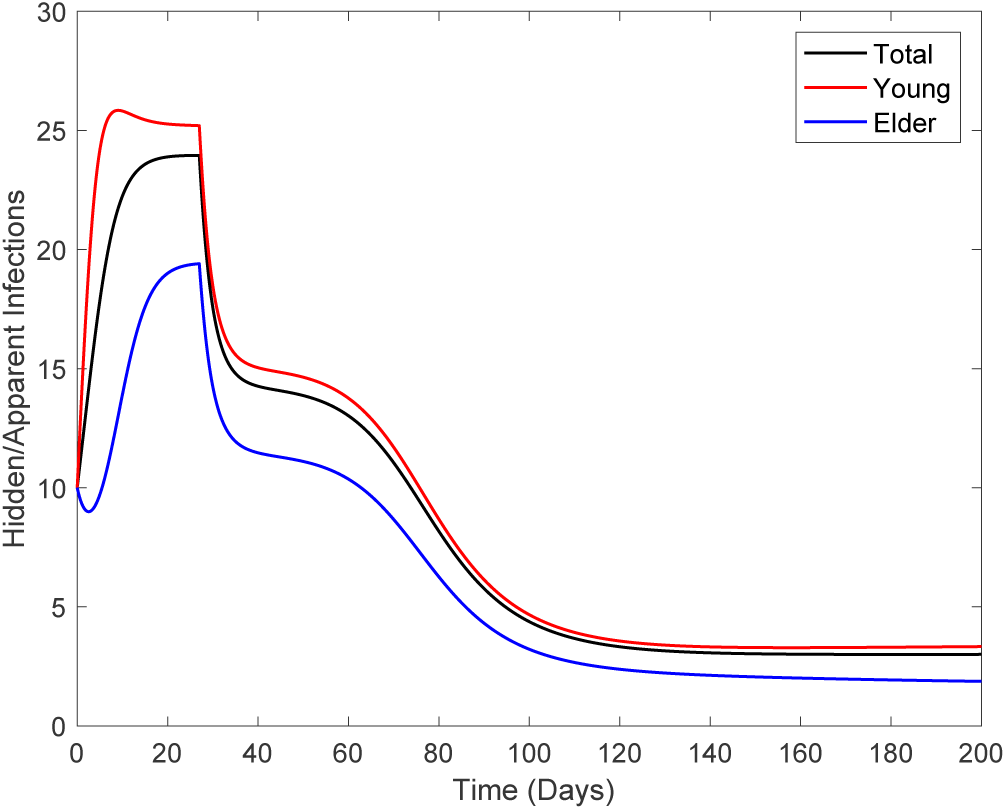
The curves of ratio hidden:apparent for young, elder and total persons.

Comparing Figures 20 and 21, as the epidemics evolves, the ratio increases quickly in the beginning, reaches a plateau during the increasing phase, and decreases during the declining phase, finally reaching another plateau after the ending phase of the first wave. In the first plateau, the ratios are 18 : 1, 26 : 1, and 24 : 1 for, respectively, elder, young, and total persons. The second plateau (5 : 1) is reached when the first wave of epidemics is ending. Therefore, during epidemics, there are much more hidden than apparent persons, which makes any control mechanisms hard if mass testing could not be implemented. However, the estimation of the ratio between hidden and apparent cases can be helpful in designing mass testing to isolated asymptomatic persons.

In this paper, epidemiological scenarios were obtained aiming the evaluation of isolation and subsequent release. However, from Figure 6(a), we observed a lowering in severe CoViD-19 data with respect to the estimated curve. Hence, let us assume that at *t =* 45, 18 days after the beginning of isolation, protection actions were adopted by persons (use of alcohol gel and face mask) [6]. Figure 22(a) shows three curves of *D*_2 =_ *D*_2_*_y_* + *D*_2_*_o_* and observed data, where 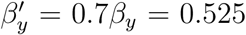, 0.5*β_y =_* 0.375, 0.45*β_y =_* 0.3375 and 0.4*β_y =_* 0.3. The better estimated is 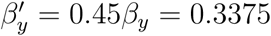. Figure 22(b) shows the curves of *D*_2_ for 4 different decreasing values of transmission rates extended from *t =* 0 until 250. We stress the fact that this evidence must be confronted with more data.

**Figure 22:**
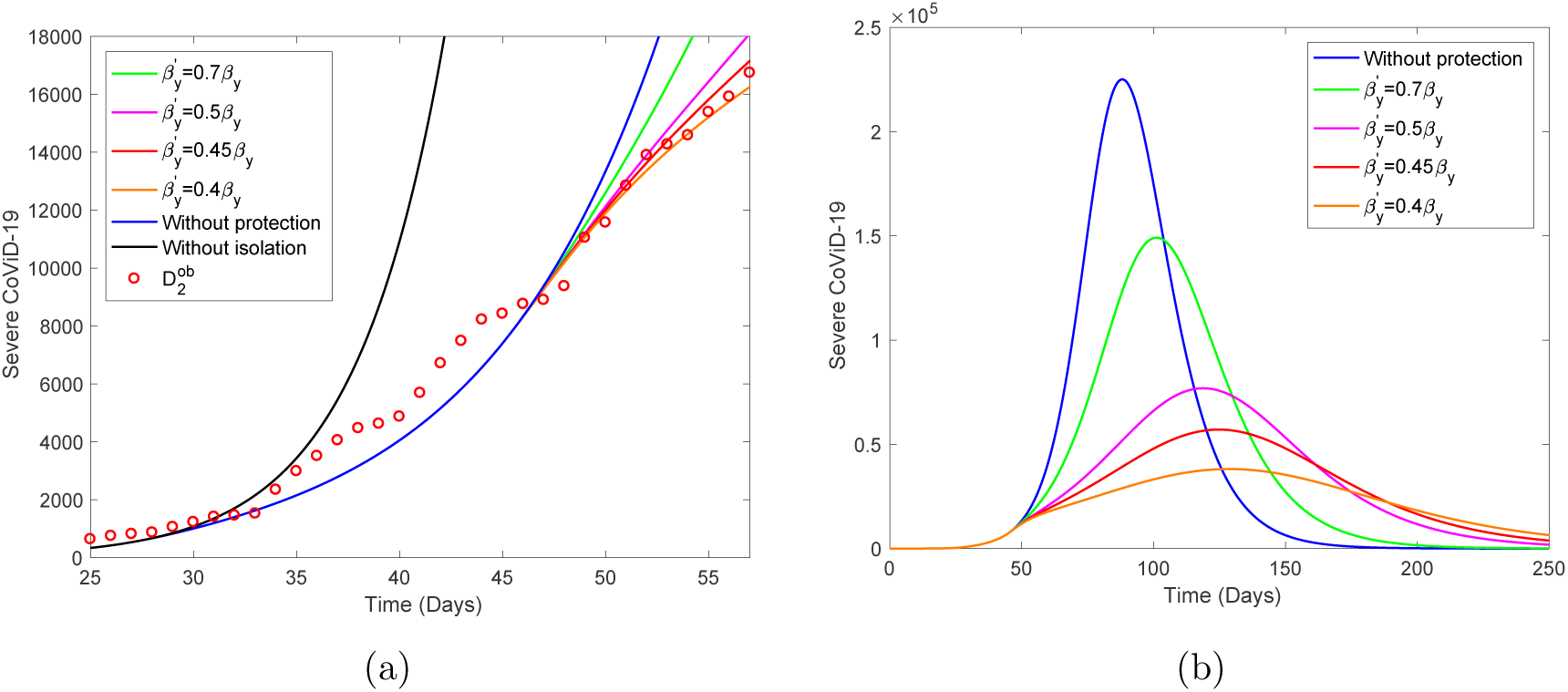
Three curves of *D*_2_ = *D*_2_*_y_* + *D*_2_*_o_* and observed data, where 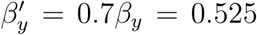, 0.45*β_y =_* 0.3375 and 0.4*β_y =_* 0.3 (a). The better estimated is 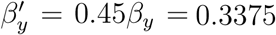. Curves of *D*_2_ for 4 different decreasing values of transmission rates extended from *t =* 0 until 250 (b)

The question of using a face mask and constant hygiene (washing hands with alcohol and gel and protection of the mouth, nose and eyes) to avoid infection is left to future work.

## 5 Conclusion

We formulated a mathematical model considering two subpopulations comprised by young and elder persons to study CoViD-19 in São Paulo State, Brazil. The model considered pulses in isolation and release. Briefly, the first CoViD-19 case was observed on February 26 (*t =* 0), isolation were initiated on March 24 (*t =* 27), and the release will be implemented on May 8 (*t =* 72).

We estimated transmission rates, additional mortality (lethality) rates, and proportions isolated from data collected in the São Paulo State. To estimate transmission rates and additional mortality (lethality) rates, we used data collected from February 26 to April 5 (*t =* 39), and data collected from March 24 to April 21 (*t =* 55), to estimate the proportion isolated. We estimated high values for the basic reproduction number, *R*_0 =_ 6.828, which is encountered among airborne viruses (see [14] for an estimation of *R*_0 =_ 6.71 for rubella infection). Currently, some authors found SARS-CoV-2 aerosol in some areas of the hospital environment, indicating the possibility of infection through air [22].

Using the estimated parameters, the system of equations (2), (3), and (4) were simulated to providing epidemiological scenarios when release will be implemented after initial isolation. Using the estimated proportion *k =* 0.5 of isolated persons, we observed that in all three strategies of releasing for regime 1 (equal release of young and elder persons), the severe CoViD-19 cases approach those without isolation, but the peaks are delayed. However, for regime 2 (releasing young but maintaining elder persons), deaths due to CoViD-19 are reduced by half among elder persons, and by 30% in all persons. Nevertheless, simulation showed that an isolation of 80% is desirable aiming reduction in the severe CoViD-19 cases.

In our model, we did not allow severe CoViD-19 cases to transmit the infection, as well as persons with mild symptoms also. This isolation assumption may have an indirect effect on the transmission of disease by two hypotheses: (1) if a virulent strain is causing severe cases, then their isolation must decrease its fitness, and (2) if severe cases release more amount of virus, then their isolation may decrease the abundance of virus in the environment. As a consequent, asymptomatic and mild cases of CoViD-19 can prevail as epidemics evolves. However, immediate consequent will be infection with more virulent strain and more absorption of the virus by heath care workers, which is a preeminent reason to provide them with extremely secure equipment.

Finally, severe CoViD-19 data collected in the São Paulo State indicates that there is another lowering in those cases besides the diminishing resulted from isolation. We hypothesize that this decreasing could be due to protection actions, but more data are needed to confirm or deny this hypothesis.

## Data Availability

The data that support the findings of this study are openly available in Ministry of Health (Brazil) at https://covid.saude.gov.br.

## A Trivial equilibrium and its stability

By the fact that *N* is varying, the system is non-autonomous non-linear differential equations. To obtain autonomous system of equations we let *k_j =_ l_ij =_* 0, *j = y, o*, and use fractions of individuals in each compartment, defined by, with *j = y* and *o*,

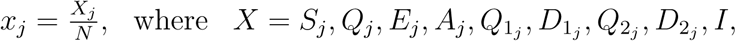

resulting in

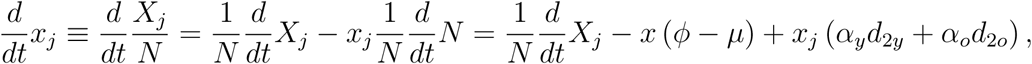

using equation (5) for *N*. Hence, equations (2), (3) and (4) in terms of fractions become, for susceptible persons,

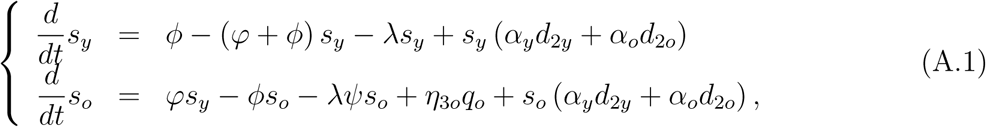

for infected persons,

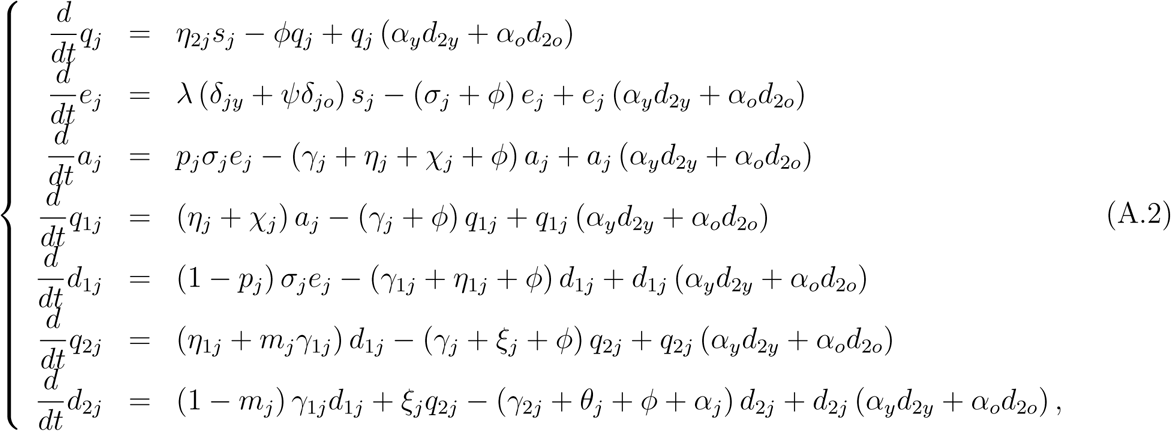

and for immune persons

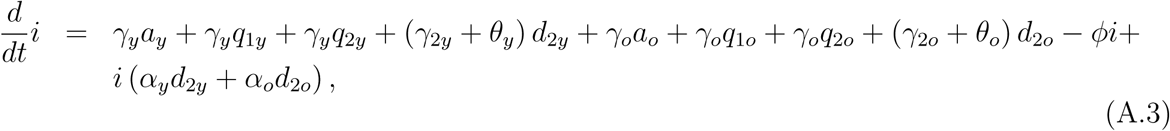

where *λ* is the force of infection given by equation (1) re-written as

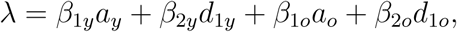

and

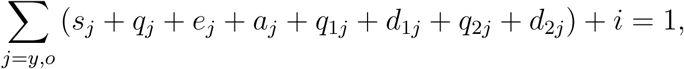

which is autonomous system of equations. We remember that all classes vary with time, however their fractions attain steady state (the sum of derivatives of all classes is zero). This system of equations is not easy to determine non-trivial (endemic) equilibrium point *P^*^*. Hence, we restrict our analysis with respect to trivial (disease free) equilibrium point.

The trivial or disease free equilibrium *P*^0^ is given by

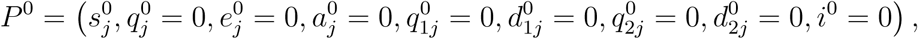

for *j = y* and *o*, where

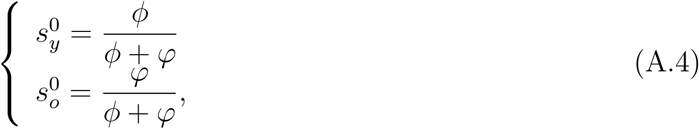

with 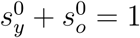.

Due to 17 equations, we do not deal with characteristic equation corresponding to Jacobian matrix evaluated at *P*^0^, but we apply the next generation matrix theory [4].

The next generation matrix, evaluated at the trivial equilibrium *P*^0^, is obtained considering the vector of variables *x =* (*e_y_,a_y_,d*_1_*_y_,e_o_,a_o_,d*_1_*_o_*). We apply method proposed in [15] and proved in [16]. There are control mechanisms (isolation), hence we obtain the reduced reproduction number *R_r_* by isolation.

In order to obtain the reduced reproduction number, diagonal matrix *V* is considered. Hence, the vectors *f* and *v* are

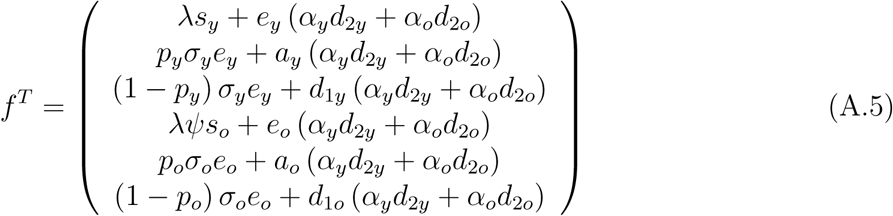

and

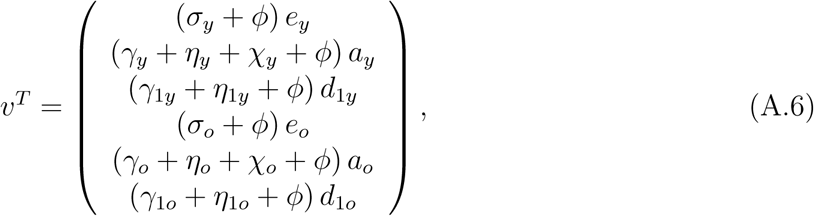

where the superscript *T* stands for the transposition of a matrix, from which we obtain the matrices *F* and *V* (see [4]) evaluated at the trivial equilibrium *P*^0^, which were omitted. The next generation matrix *FV*^−1^ is

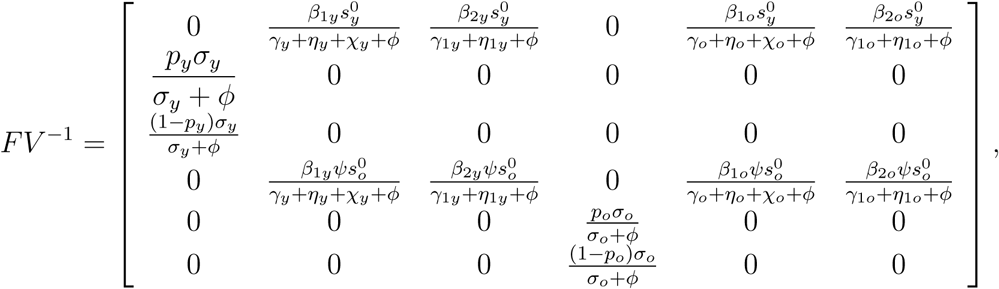

and the characteristic equation corresponding to *FV*^−1^ is

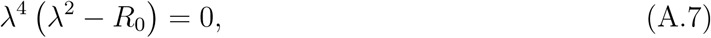

where the basic reproduction number *R*_0_ is

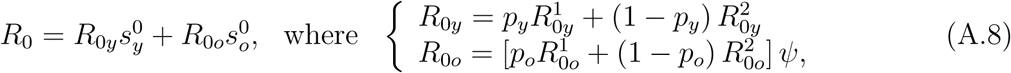

and *R*_0_*_y_* and *R*_0_*_o_* are the basic partial reproduction numbers defined by

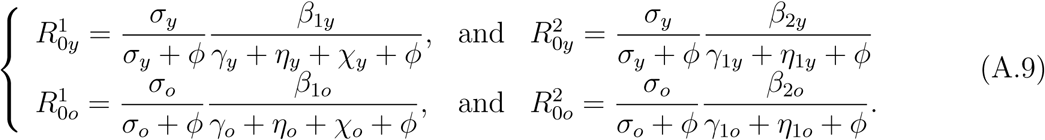

Instead of using the spectral radius 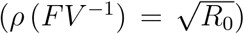, we apply procedure in [15] (the sum of coefficients of characteristic equation), resulting in a threshold *R*_0_. Hence, the trivial equilibrium point *P*^0^ is locally asymptotically stable (LAS) if *R*_0_ *<* 1.

In order to obtain the fractions of susceptible individuals, *M* must be the simplest (matrix with least number of non-zeros). Hence, the vectors *f* and *v* are

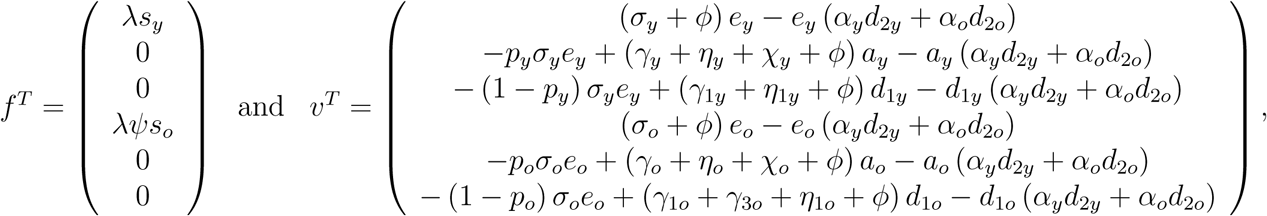

where superscript *T* stands for the transposition of a matrix, from which we obtain the matrices *F* and *V* evaluated at the trivial equilibrium *P*^0^, which were omitted. The next generation matrix *FV*^−1^ is

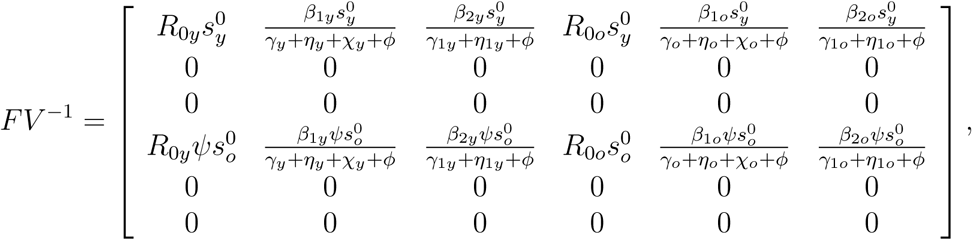

and the characteristic equation corresponding to *FV*^−1^ is

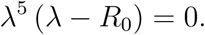

The spectral radius is 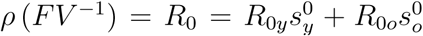 given by equation (A.8). Hence, the trivial equilibrium point *P*^0^ is LAS if *ρ <* 1.

Both procedures resulted in the same threshold, hence, according to [20], the inverse of the reduced reproduction number *R*_0_ given by equation (A.8) is a function of the fraction of susceptible individuals at endemic equilibrium *s*^*^ through

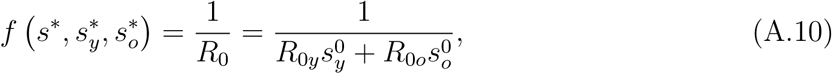

where 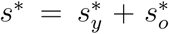 (see [18] [20]). For this reason, the effective reproduction number *R_ef_* [17], which varies with time, can not be defined neither by *R_ef =_ R*_0_ (*s_y_* + *s_o_*), nor *R_ef =_ R*_0_*_y_s_y_* + *R*_0_*_o_s_o_*. The function *f* (ϰ) is determined by calculating the coordinates of the non-trivial equilibrium point *P*^*^. For instance, for dengue transmission model, 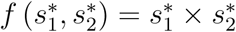, where 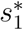 and 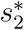 are the fractions at equilibrium of, respectively, humans and mosquitoes [18]. For tuberculosis model considering drug-sensitive and resistant strains, there is not *f* (ϰ), but *s*^*^ is solution of a second degree polynomial [20].

From equation (A.10), let us assume that 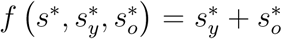. Then, we can define the approximated effective reproduction number *R_ef_* as

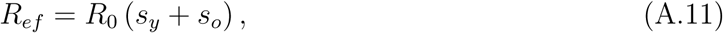

which depends on time, and when attains steady state (*R_ef =_* 1), we have *s*^*^ *=* 1/*R*_0_.

The basic partial reproduction number 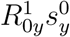 (or 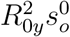) is the secondary cases produced by one case of asymptomatic individual (or pre-diseased individual) in a completely susceptible young persons without control; and the partial basic reproduction number 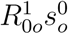 (or 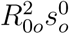) is the secondary cases produced by one case of asymptomatic individual (or pre-diseased individual) in a completely susceptible elder persons without control. If all parameters are equal, and *ψ =* 1, then

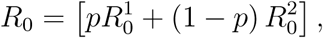

where 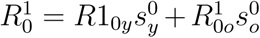 and 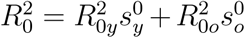 are the basic partial reproduction numbers due to asymptomatic and pre-diseased persons.

The global stability follows method proposed in [9]. Let the vector of variables be *x =* (*e_y_,a_y_,d*_1_*_y_,e_o_,a_o_,d*_1_*_o_*), vectors *f* and *v*, by equations (A.5) and (A.6), and matrices *F* and *V* evaluated from *f* and *v* at trivial equilibrium *P*^0^ (omitted here). Vector *g*, constructed as

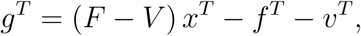

results in

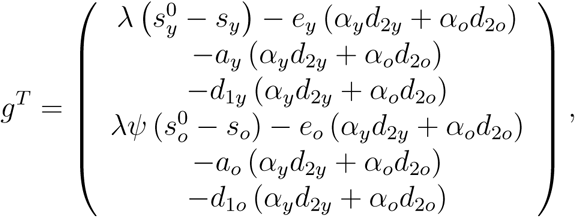

and *g^T^* ≥ 0 if 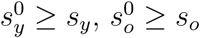 and *α_y =_ α_o =_* 0.

Let *v_l =_* (*z*_1_*,z*_2_*,z*_3_*,z*_4_*,z*_5_*,z*_6_) be the left eigenvector satisfying *v_l_V*^−1^ *F = ρv_l_*, where 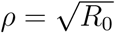,
and

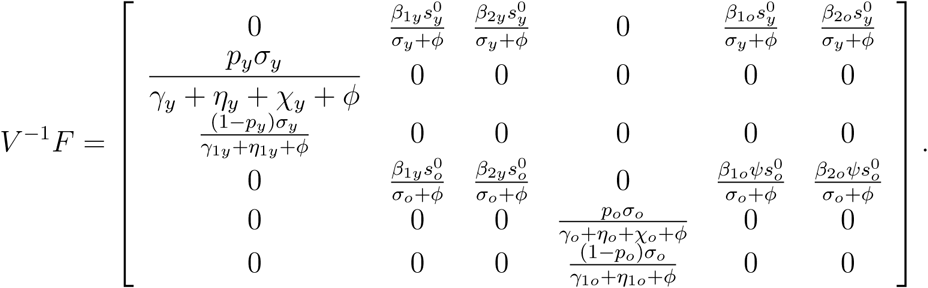

This vector is

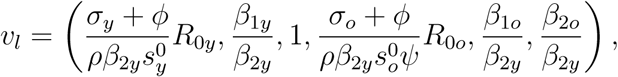

and Lyapunov function *L*, constructed as *L = v_l_V*^−1^*x_T,_* is

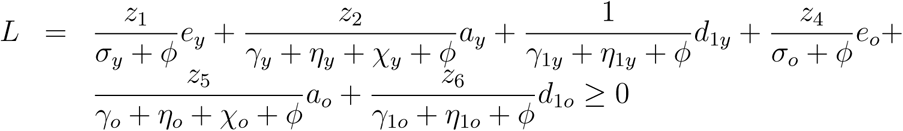

always, and

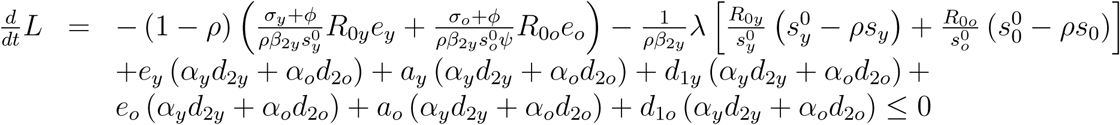

only if 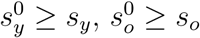 and *α_y =_ α_o =_* 0.

Hence, the method proposed in [9] is valid only for α*_y =_ α_o =_* 0, in which case *P*^0^ is globally stable if 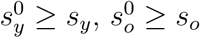 and 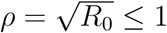.

## Footnotes

1 Initially, programmed to end on April 6, further extended to April 22, and postponed to May 10.

2 Simulations were done on April 21. We anticipated the release to May 8 (May 10 is Mothers’ day).

## References

[1] Adesão ao isolamento social em SP (April 27, 2020). https://www.saopaulo.sp.gov.br/coronavirus/isolamento.

[2] R.M. Anderson, R.M. May, R.M., Infectious Diseases of Human. Dynamics and Control, Oxford University Press, Oxford, New York, Tokyo (1991).

[3] Boletim Epidemiológico 08 (April 9, 2020), https://www.saude.gov.br/images/pdf/2020/April/09/be-covid-08-final-2.pdf.

[4] O. Diekmann, J.A.P. Heesterbeek, M.G. Roberts, The construction of next-generation matrices for compartmental epidemic models, J. R. Soc. Interface 7 (2010) 873–885.

[5] N.M. Ferguson, et al., Impact of non-pharmaceutical interventions (NPIs) to reduce COVID-19 mortality and healthcare demand, Imperial College COVID-19 Response Team (2020).

[6] J. Howad, et al., Face masks against COVID-19: An evidence review, (April 12, 2020), doi:10.20944/preprints202004.0203.v1.

[7] S.M. Raimundo, H.M. Yang, R.C. Bassanezi, M.A.C. Ferreira, The attracting basins and the assessment of the transmission coefficients for HIV and M. tuberculosis infections among women inmates, Journal of Biological Systems (10)(1) (2002) 61–83.

[8] SEADE—Fundação Sistema Estadual, https://www.seade.gov.br (2020).

[9] Z. Shuai, P. van den Driessche, Global stability of infectious disease model using Lyapunov functions, SIAM J. App. Math. 73(4) (2013) 1513–1532.

[10] WHO, Report of the WHO-China Joint Mission on Coronavirus Disease 2019 (COVID-19), 16–24 February 2020 (2020).

[11] H.M. Yang, Modelling vaccination strategy against directly transmitted diseases using a series of pulses, Journal of Biological Systems (6)(2) (1998) 187–212.

[12] H.M. Yang, Directly transmitted infections modeling considering age-structured contact rate—Epidemiological analysis, Mathematical and Computer Modelling 29(7) (1999) 11–30.

[13] H.M. Yang, Directly transmitted infections modeling considering age-structured contact rate, Mathematical and Computer Modelling, 29(8) (1999) 39–48.

[14] H.M. Yang, Modeling directly transmitted infections in a routinely vaccinated population—The force of infection described by Volterra integral equation, Applied Mathematics and Computation (122)(1) (2001) 27–58.

[15] H.M. Yang, The basic reproduction number obtained from Jacobian and next generation matrices—A case study of dengue transmission modelling, BioSystems 126 (2014) 52–75.

[16] H.M. Yang, D. Greenhalgh, Proof of conjecture in: The basic reproduction number obtained from Jacobian and next generation matrices—A case study of dengue transmission modelling, Appl. Math. Comput. 265 (2015) 103–107.

[17] H.M. Yang, J.L. Boldrini, A.C. Fassoni, K.K.B. Lima, L.S.F. Freitas, M.C. Gomez, V.F. Andrade, A.R.R. Freitas, Fitting the incidence data from the City of Campinas, Brazil, based on dengue transmission modellings considering time-dependent entomological parameters, PlosOne (March 24) (2016) 1–41.

[18] H.M. Yang, The transovarial transmission un the dynamics of dengue infection: Epidemiological implications and thresholds, Math. Biosc. 286 (2017) 1–15.

[19] M.C. Gomez, H.M. Yang, A simple mathematical model to describe antibody-dependent enhancement in heterologous secondary infection in dengue, Mathem. Med. Biol.: A Journal of the IMA?? (2018), 1–28.

[20] H.M. Yang, Are the beginning and ending phases of epidemics provided by next generation matrices?—Revisiting drug sensitive and resistant tuberculosis model, Appl. Math. Comput., submitted (2020).

[21] H.M. Yang, Modeling the transmission of new coronavirus in the São Paulo State, Brazil—Assessing epidemiological impacts of isolating young and elder persons, Mathem. Med. and Biol.: A journal of the IMA, submitted (2020).

[22] L. Yuan, et al., Aerodynamic analysis of SARS-CoV-2 in two Wuhan hospitals, https://doi.org/10.1038/s41586-020-2271-3 (2020).

